# IL-32 producing CD8^+^ memory T cells and Tregs define the IDO1 / PD-L1 niche in human cutaneous leishmaniasis skin lesions

**DOI:** 10.1101/2024.01.02.23300281

**Authors:** Nidhi S. Dey, Shoumit Dey, Naj Brown, Sujai Senarathne, Luiza Campos Reis, Ritika Sengupta, Jose Angelo L. Lindoso, Sally James, Lesley Gilbert, Mitali Chatterjee, Hiro Goto, Shalindra Ranasinghe, Paul M. Kaye

## Abstract

Human cutaneous leishmaniasis (CL) is characterised by chronic skin pathology. Experimental and clinical data suggest that immune checkpoints (ICs) play a crucial role in disease outcome but the cellular and molecular niches that facilitate IC expression during leishmaniasis are ill-defined. We previously showed that in Sri Lankan patients with CL, indoleamine 2,3-dioxygenase 1 (IDO1) and programmed death-ligand 1 (PD-L1) are enriched in lesion skin and that reduced PD-L1 expression early after treatment onset predicted cure rate following antimonial therapy. Here, we use spatial cell interaction mapping to identify IL-32-expressing CD8^+^ memory cells and regulatory T cells as key components of the IDO1 / PD-L1 niche in Sri Lankan CL patients and in patients with distinct forms of dermal leishmaniasis in Brazil and India. Furthermore, the abundance of IL-32^+^ cells and IL-32^+^CD8^+^ T cells at treatment onset was prognostic for rate of cure in Sri Lankan patients. This study provides a unique spatial perspective on the mechanisms underpinning IC expression during CL and a novel route to identify additional biomarkers of treatment response.

**Graphical Abstract:** 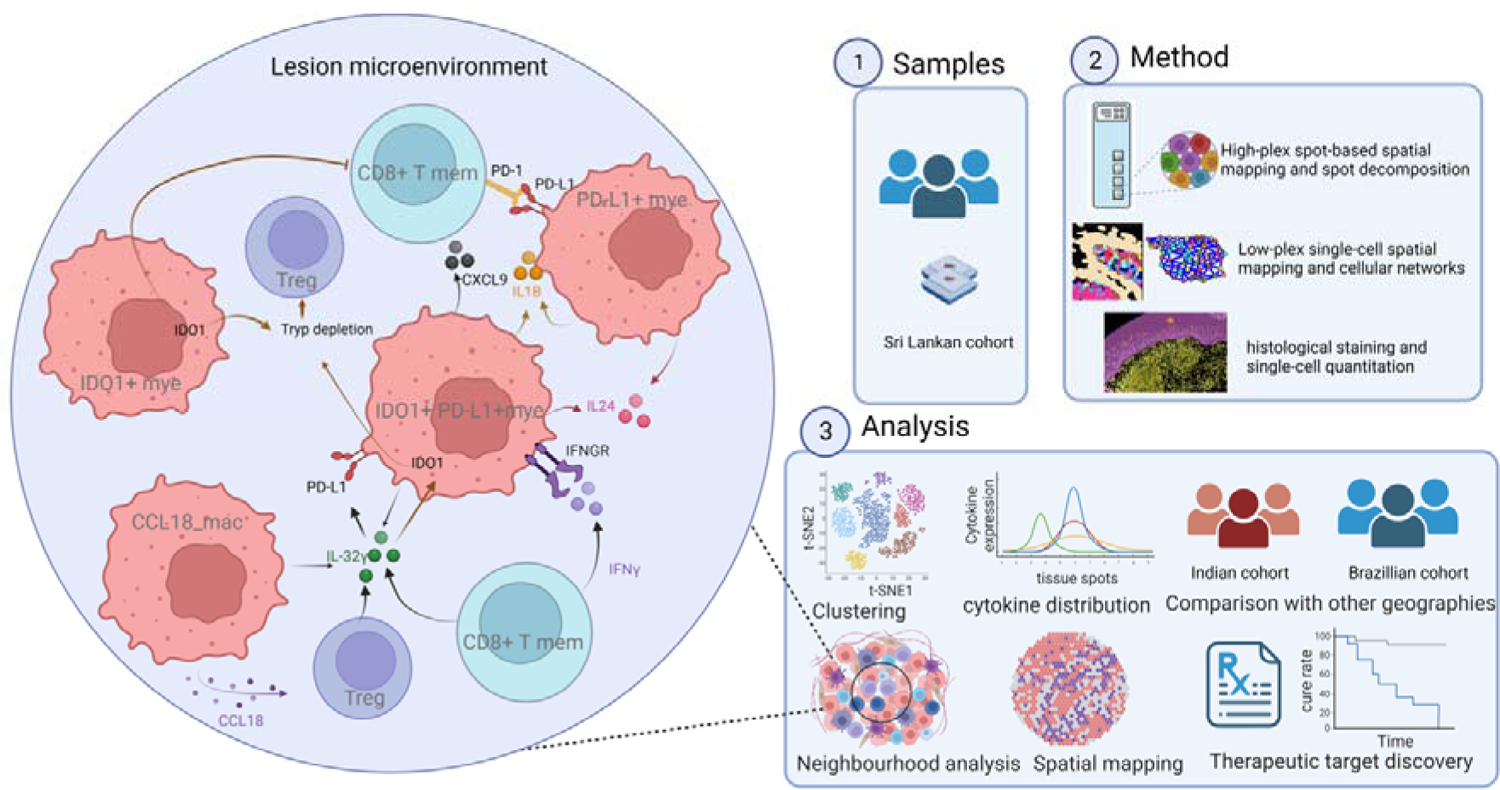

## Introduction

Cutaneous leishmaniasis (CL) is caused by protozoan parasites of the genus *Leishmania* and exhibits a wide spectrum of clinical presentations determined at least in part by the host immune response. *Leishmania* are intracellular parasites of myeloid cells, predominantly macrophages, monocytes, and dendritic cells ^1, 2, 3^. Consequently, protective immunity is largely cell-mediated, with effector CD4^+^ and CD8^+^ T cells producing cytokines (e.g. IFNγ) that activate myeloid cell-intrinsic leishmanicidal activity ^4, 5^. Over-exuberant or persistent effector T cell function at the site of infection can lead to tissue damage and progression of clinical disease despite a reduction in parasite load ^6^. Conversely, parasite persistence has been attributed to a combination of dysregulation of the T cell response and parasite-mediated subversion of epigenetic and transcriptional pathways that control macrophage polarisation and activation ^7^.

Studies in animal models have elegantly demonstrated the impact of dichotomy in CD4^+^ T cell responses to *Leishmania*. For example, enhanced CD4^+^ Th2 responses ^8^ are responsible for the genetically determined susceptibility of BALB/c mice to *L. major* infection and IL-4-driven alternatively activated macrophages favour parasite survival ^9^. This dichotomy in CD4^+^ T cell response has some parallels in human leishmaniasis. Strong Th1 responses are associated with resolution of infection in CL patients infected with *L. donovani* in Sri Lanka ^9^ and with *L. major* in Iran ^10^, whereas increased lesional IL-4 was observed in Sri Lankan patients with a poor response to treatment ^11^. However, chronic stimulation of CD4^+^ Th1 cells leads to differentiation into Tr1 cells producing IL-10, a well-known inhibitor of IFNγ-mediated NO production ^12^. Regulatory T cells producing IL-10 have also been associated with parasite latency, treatment resistance, and disease relapse in humans ^13, 14^. In *L. braziliensis* infection, chronic lesion development occurs in the face of relatively low parasite burden and has been most commonly associated with an over-exuberant cytolytic CD8^+^ T cell response ^15^.

In leishmaniasis ^16^ as well as many other infectious ^17^ and non-infectious ^18, 19^ diseases, the role of immune checkpoints (ICs) as regulators of T cell effector function and disease outcome has been well described. For example, indoleamine-2,3-dioxygenase (IDO1) is a metabolic IC molecule expressed in response to inflammatory insults by macrophages ^20^, dendritic cells ^21^ and B cells ^22^ and catalyses degradation of tryptophan ^23^. Tryptophan starvation in T cells leads to proliferation arrest ^23^, anergy ^24^ or regulatory phenotypes ^25^, facilitating tumour immune escape ^26^ or in the case of CL lesion progression ^27^. Similarly, programmed death ligand 1 (PD-L1; *CD274*) is expressed by myeloid cells upon activation via LPS/IFN-γ or IL-4 and when bound to PD-1 on T cells inhibits activation and promotes IL-10 expression ^28^. These and other ICs have become invaluable prognostic and therapeutic targets in cancer. Co-expression of PD-L1 and IDO1 in resected non-small cell lung cancer (NSCLC) may predict survival outcome ^29^ and triple blockade of IDO1, PD-L1 and MAPK/ERK kinase has been proposed as therapy in NSCLC ^30^. In a similar vein, an immunomodulatory vaccine targeting IDO1^+^PD-L1^+^ myeloid cells is under development for metastatic melanoma ^31^. In addition to modulating the natural course of disease, ICs may also play a role in tempering the efficacy of conventional but immune-dependent anti-infectives and anti-cancer drugs. For example, we recently demonstrated that PD-L1 and IDO1 are enriched in skin lesions of CL patients in Sri Lanka and that a decrease in PD-L1 expression early after treatment onset was predictive of rate of cure following treatment with sodium stibogluconate, a well-known immune dependent drug ^32^.

Despite many examples of disease-associated aberrant expression of IDO1 and PD-L1^33, 34, 35, 36, 37, 38^, our understanding of the cellular and molecular pathways that regulate expression of the ICs is largely derived from in vitro studies. Multiple cytokines and inflammatory signals can induce IDO1 ^39, 40^ and PD-L1 ^41, 42^ expression on human monocytes and DCs, including IFNγ, TNF, TGF-β, IL-6, IL-10, IL-27, IL-32, PAMPs/DAMPs, and PGE2. IDO1 and PD-L1 are also induced by intracellular infection with *Leishmania* ^32, 43, 44^ and other pathogens ^45, 46^ suggesting that subversion of host cell function by manipulation of ICs is a conserved mechanism across pathogen evolution. There is currently little understanding, however, of how these ICs are regulated in the complex spatial microenvironments associated with chronic tissue pathology that result from either infection or cancer.

Here, we have combined multiple spatial methodologies to define the cellular and transcriptomic composition of niches containing IDO1^+^ and PD-L1^+^ myeloid cells in tissue biopsies from Sri Lankan CL patients and patients with other diverse forms of dermal leishmaniasis. Our results reveal diversity in niche composition and localisation, but identify IL-32^+^ CD8^+^ T cells and IL-32^+^ Tregs as common attributes of these immunoregulatory microenvironments.

## Results

### *CD274* and *IDO1* localise to myeloid cell-rich niches

To understand the cellular and molecular architecture of human CL lesions, we conducted Visium spatial transcriptomics on FFPE sections from 6 patients (P1-P6; **Supplementary Table 1**) presenting with a papular and/or ulcerative plaque lesion typical of Sri Lankan CL (SL_CL) (**Fig.1a** and **Extended Data Fig.1a**). H&E staining and immunochemistry revealed dense cellular infiltration and parasitism in the papillary dermis (**Fig.1b** and **Extended Data Fig.1b**). A total of 2418 Visium spots (median of 4104 gene counts per spot; **Extended Data Fig.1c**) were coloured by cluster identities and visualised in UMAP space, with representation from all 6 patients in each cluster (**Fig.1c**). 11 high level clusters were identified and annotated namely My1, My2 and My3 (Myeloid rich), B/Fib (cluster rich in B and Fibroblast transcripts), TL (T lymphocyte rich), KC1 (Keratinocyte 1 rich), Fib (Fibroblast 1 rich), KC2 (Keratinocyte 2 rich), B (B cell rich), and Endo (Endothelial cell rich) and an uncharacterised cluster (mix) (**Fig. 1d,e**). Spatial mapping generated a coarse transcriptomic map reflecting underlying tissue morphology (**Fig. 1f** and **Extended Data Fig. 1b,d**). The three myeloid rich clusters had distinct positioning and gene expression signatures. My1 (*SELENOP*+) and My2 (*CCL18*^+^) were located in the papillary dermis near the epidermal-dermal junction (herein referred to as “lesion core”), My1 also contained B cells (*IGHA1*, *IGHG2*, *IGKC*) (**Supplementary** Fig. 1a). My1 and My2 were enriched for mRNAs encoding i) S100 proteins (*S100A8*, *S100A9*) suggestive of neutrophils, monocytes and DC^47^; ii) metallothionein genes (*MT1H, MT1G, MT1X* and *MT2A*) and *SLC39A8*, suggesting altered metal ion homeostasis; and iii) *CCL18*, a T cell chemoattractant ^48^ (**Supplementary** Fig. 1b). My3 (*CHIT1*^+^) was associated with a T lymphocyte-rich cluster (*CCL19*^+^) deeper in the dermis (**Extended Data Fig. 1d**). *IDO1* and *CD274* transcripts mapped to the lesion core and to the T cell rich region (**Fig. 1g,h**, and **Supplementary** Fig. 2a,b) and to the My1, My2 and My3 clusters (**Fig. 1i**). My2 was also enriched for other immune regulators including *SIGLEC10*^49^, *VSIG4*^50^ and *CD300E*^51^ (**Supplementary** Fig. 1b).

**Fig. 1:**
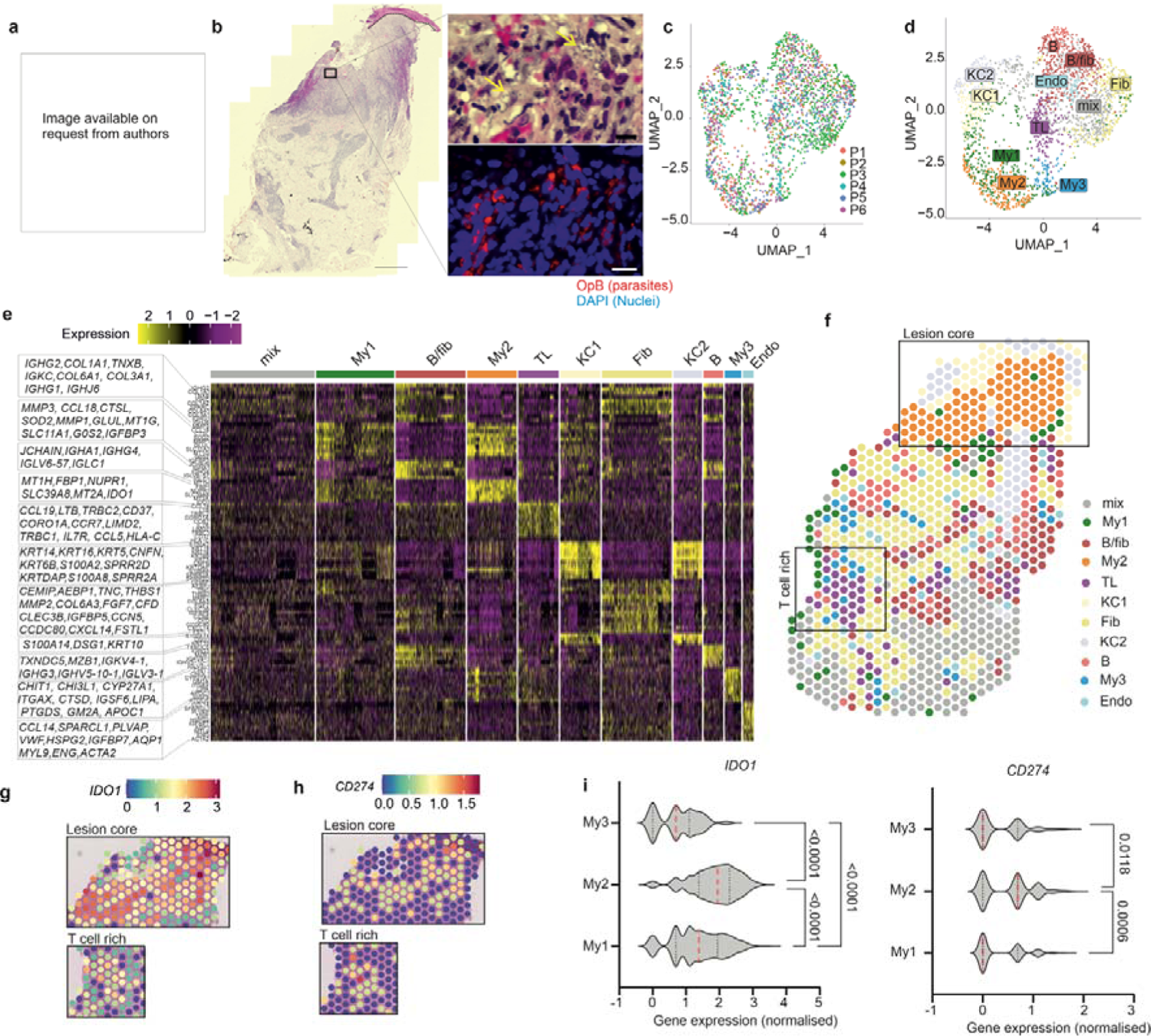
Spatial transcriptomics identifies spatial domains in *L. donovani* CL lesions. **a**, Patient P3 showing lesion macroscopic features. **b,** Left, histology of P3 shows dense dermal infiltrate (Hematoxylin and eosin; H & E; Bar 500µm). Right, Higher magnification of site of parasitism; top, H&E and bottom, anti-*Leishmania* OPB staining. Bar 10 µm. Parasites indicated in H&E by yellow arrow. **c,** UMAP projection of Visium spots from patients P1-P6 coloured by patient ID. **d,** as (**c**) but clustered, coloured and labelled by gene expression. **e,** Heatmap showing the top 10 genes enriched in each cluster. **f,** Spatial map for patient P3 coloured by clusters identified in (**d)**. **g-h,** Spatial feature plots for P3 showing normalised values of *CD274* (**g**) and *IDO1* (**h**) **i,** *IDO1* and *CD274* violin plots for My1-3 clusters for P1-6(red dotted line, mean; black dotted lines, IC range) p-values for group mean ranks compared by Kruskal-Wallis one-way test.

Using a publicly available scRNAseq dataset^38^ and the Cell2Location^52^ prediction tool, we projected estimated abundances of 40 cell types (**Extended Data Fig. 2**). Although some interpatient heterogeneity was observed, macrophages (*FCGR2A*, *F13A1*, *NR4A1*, *NR4A2*, *KLF4*), helper T cells (*CD4*, *IL7R*, *CD40LG*, *PTGER4*), regulatory T cells (*FOXP3*, *TIGIT*, *BATF*, *CTLA4*), natural killer cells (*KLRD1*, *GNLY*, *PRF1*, *GZMB*, *NKG7*), and cytotoxic T cells (*CD8A*, *CD8B*) were most highly represented. (**Extended Data Fig. 2a**). My1, My2 and My3 clusters contained Tc, Th, Treg and NK cells, myeloid DC (DC2; *CD68, NR4A1, NR4A2, CLEC10A, FCGR2A, CD83^low^*), ILC2 (*IL7R, PTGDR2, GATA3*), Macro1(*MARCO*, *CD163, C1QB, FCGR2A*), Macro2, monocytes (*CD14*, *1L1B*) and ILC1/NK (*KLRB1, XCL1, XCL2, TNFRSF18, TNFRSF11, FCER1G, KIT*) in differing proportions (**Extended Data** Fig. 2b-l). Although Ig transcripts were found in these clusters, formal identification of B cells / plasma cells was not possible using the Reynolds et al reference data set^38^.

### Mature dendritic cells express abundant *CD274* and *IDO1* mRNA

To overcome limitations of cellular deconvolution, we next applied single cell spatial transcriptomics. Using Nanostring CosMx^53^, we analyzed 115,157 single cells from 20 fields of view (FOVs) derived from four patients (**P3-P6**; **Extended Data** Fig. 3a-e) (**Supplementary Table 1**) and identified 22 cell clusters (**Fig.2a** and **Extended Data** Fig. 3f,g). Cells with myeloid signatures were most abundant in the papillary dermis (**Fig. 2b**), supporting our Visium analysis. We also identified T cells near the dermal and epidermal boundary, flanked by fibroblasts and B cells in the lesion core. Deeper in the dermis, T cells along with scattered macrophages were dominant (**Fig.2b**). The localisation of additional cell populations is shown in **Extended Data** Fig. 4a-d. We next subclustered all myeloid cells (mac, mac2, mDC, pDC, monocytes, and neutrophils) to obtain 13 sub-populations (**Fig. 2c,d** and **Extended Data** Fig. 4e,h). When ranked by *CD274* and *IDO1* mRNA abundance (**Extended Data** Fig. 4i,j), mature dendritic cell cluster DC3 was identified as having the highest proportion of *IDO1*^hi^ *CD274*^hi^ cells. Smaller proportions of many other myeloid cell populations also expressed these ICs. DC3 mapped to the lesion core and the deeper T cell rich regions of the dermis (**Fig.2e**) and expressed mRNA for other immunoregulatory molecules (*CD40* and *PDCD1LG2*; PD-L2; **Fig. 2d**).

**Fig. 2:**
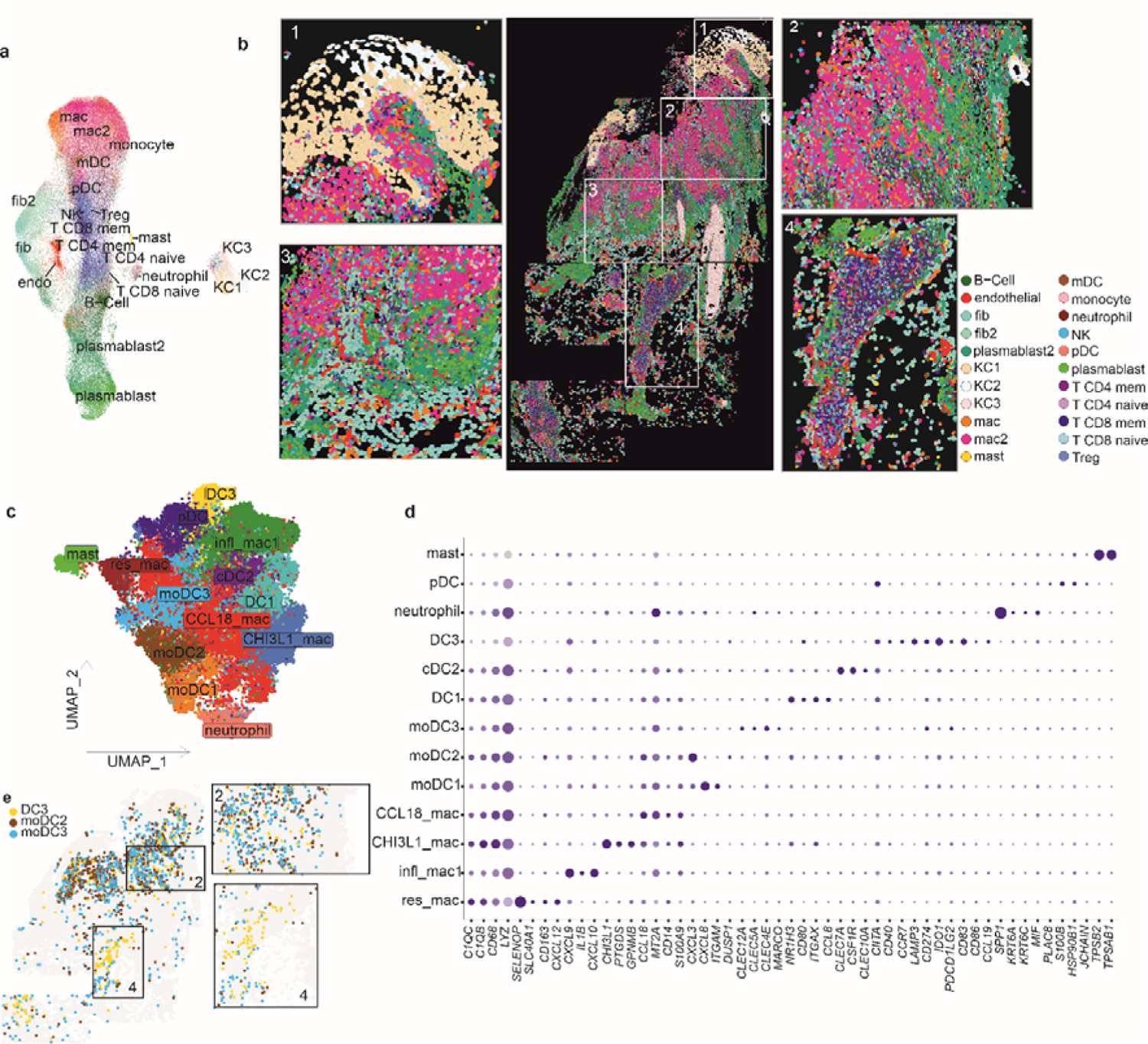
Single-cell transcriptomic imaging of cutaneous lesions from Sri Lanka. **a,b**, Single cell spatial mapping of CL lesions. **a,** UMAP representation of single cell data from 4 patients coloured by cell type **b,** single cell representation of P3 lesion biopsy generated by stitching individual FOVs. Boxes 1-4 provide higher magnification views of selected regions of interest. **c**. Same as (**a**) but for sub-clustered myeloid cells only. **d,** Dot plot showing gene expression across the sub-clustered myeloid cell types in (**c)**. **e,** localisation of DC3, moDC2 and on patient P3 with representative magnified regions of interest.

### *CD274* and *IDO1* mRNA abundance correlates with distinct cytokines and chemokines

To gain insights into the pathways leading to IDO1 and PD-L1 expression, we first sought to identify cytokine expression in niches where *CD274*^+^ and *IDO1*^+^ cells were abundant. Due to the limited cytokine / chemokine coverage of the CosMx gene panel, we reverted for this analysis to our Visium data. We classified Visium spots into four classes: *CD274*^hi^*IDO1*^lo^ (CD274 spots), *CD274*^lo^*IDO1*^hi^ (IDO1 spots), *CD274*^hi^*IDO1*^hi^ (IDO1/CD274 spots) or *CD274*^lo^ *IDO1*^lo^ (rest of the spots) and mapped their distributions and predicted cell type abundances (**Fig. 3a,b** and **Extended Data** Fig. 5a-d)). As compared to CD274 spots, IDO1/CD274 spots and IDO1 spots had a higher monocyte abundance and higher cytotoxic T cell: helper T cell ratio. We performed differential expression analysis to identify cytokines, chemokines, their receptors, and ICs that were significantly different across these classes (**Extended Data** Fig. 5e) and observed four distinct patterns of gene expression (**Fig. 3c-k** and **Extended Data** Fig. 5f,g). *CCL18, IL24*, *IL1B*, *TNFRSF6B, IFNGR2*, *CXCL9* and *IL32* were highly expressed in IDO1/CD274 spots and to a lesser degree in IDO1 spots in the lesion core, with *CXCL9* and *IL32* also found in the T cell rich hypodermis (**Fig. 3d-g**). The homeostatic chemokines *CXCL12* ^54^ and *CXCL14* ^55^ showed a reciprocal distribution being associated with spots lacking *IDO1* and *CD274* (**Fig. 3h,i**). *CCL19* (**Fig. 3j**), *CCR7*, *CXCL13* and *LTB* (**Fig. 3k**) were most highly expressed in CD274 spots in T cell rich areas. Amongst anti-leishmanial effector and regulatory cytokines ^1^, *TNF* and *IFNG* were mostly concentrated in the lesion core while *TGFB1* was widespread across the tissue (**Extended Data** Fig. 5h-j).

**Fig. 3:**
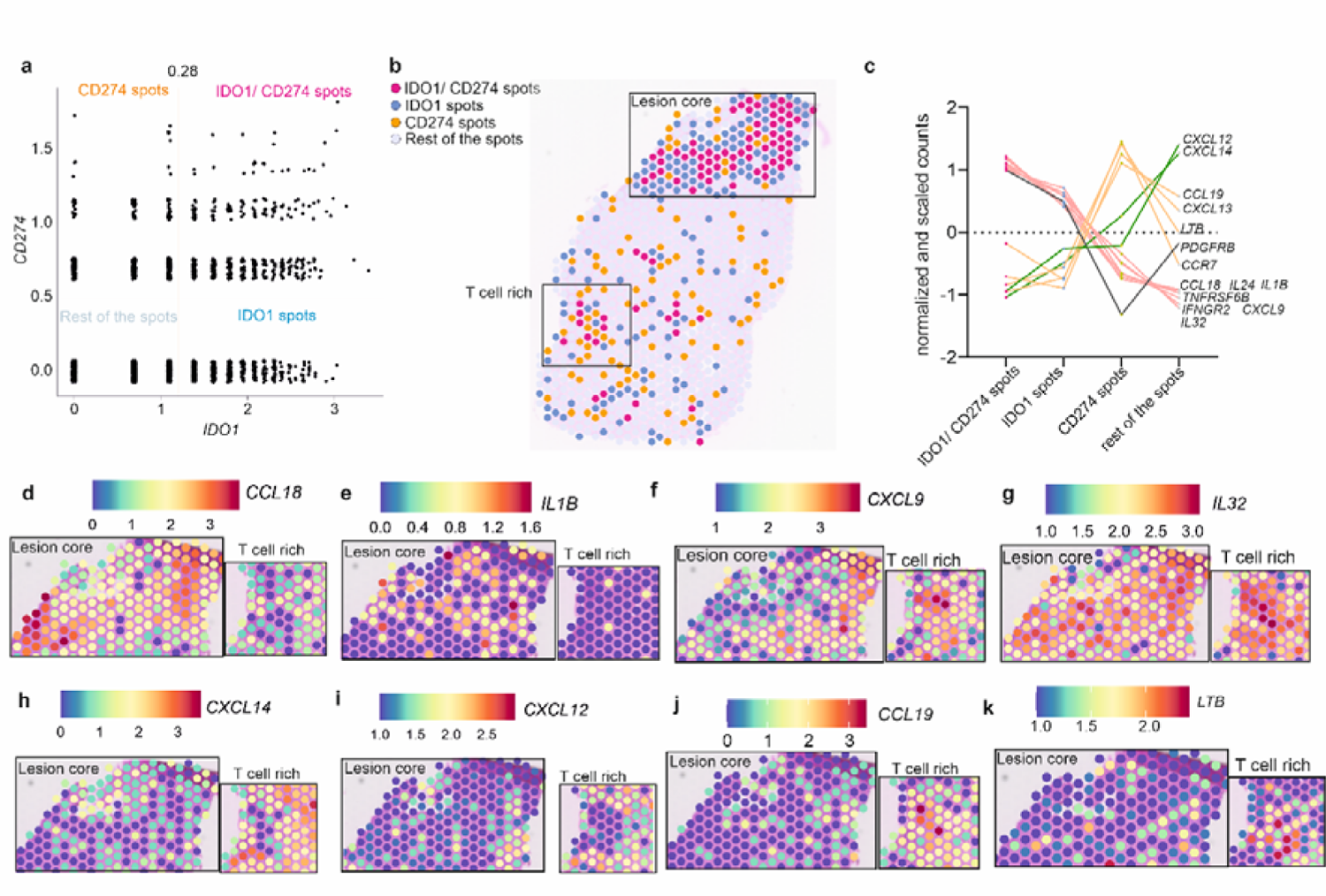
55 µm neighbourhoods of *IDO1^+^* and *CD274*^+^ cells. **a**, Scatterplot of *IDO1* and *CD274* normalised gene expression for all 55um Visium spots (for patients P3-P6). Thresholds were drawn at x=1.1 and y = 0.5 to classify spots as IDO1, CD274, IDO1/ CD274 or rest of the spots. **b**. Representative spatial plot coloured by spot class in (**a**) shown for P3. Insets identify lesion core and dermal T cell rich regions as inferred from Visium and CosMx datasets. **c**, Cytokine, chemokine, or receptor abundance by IDO1 and PD-L1 expression class described in (**a)** across n=4 patients. **d-k**, Spatial feature plots for cytokines, chemokines, and interleukins from (**c)** in lesion core and T cell rich area for patient P3.

### Neighbourhood analysis of *CD274* and *IDO1* expressing cells

To identify cellular interactions contributing to the *CD274* and *IDO1* niches, we similarly classified myeloid cells in our CosMx dataset into 4 classes by *IDO1* and *CD274* expression (**Fig. 4a,b** and **Extended Data Fig.6a,b**). CCL18^+^ macrophages (*CCL18, MT2A, CD14, S100A9, C1QB, CD68, LYZ*) were the predominant cell type across all three classes (**Fig. 4c** and **Extended Data 6a,b**). We then used Delaunay triangulation in Giotto to construct a pan spatial network based on cell centroid physical distances ^56^. We observed that each cell had 4-8 close “neighbours” (**Extended Data** Fig. 6c,d). Using this framework, we assigned IDO1^+^mye, CD274^+^mye and IDO1^+^/ CD274^+^mye cells as ‘source’ cells or as “both” neighbour and source (**Fig.4d, e** and **Extended Data Fig.6e,f**) and visualised source-neighbour or both-neighbour abundance using the UpSetR tool ^57^ (**Fig.4f** and **Extended Data Fig. 6 g,h**). This analysis revealed a consistent neighbourhood composition, comprising other *IDO1*^+^ or *CD274*^+^ myeloid cells, *CD274*^-^*IDO1*^-^ *CCL18*^+^ macrophages, CD8 memory T cells and regulatory T cells (Tregs) (**Fig. 4g-i**). For orthogonal validation, we performed immunostaining for PD-L1, IDO1, and CD8 proteins in biopsies where sufficient tissue was available (n=23 SL-CL patients), demonstrating CD8^+^ T cells in close proximity to IDO1 and PD-L1 positive cells (**Fig. 4j**). Employing bespoke image analysis pipelines, we created image masks at distances of 25, 50, and 100 microns from IDO1^+^, PD-L1^+^ (**Extended Data Fig. 6i,j**) and IDO1^+^ PD-L1^+^ cells (**Fig. 4k**), revealing that the majority of CD8^+^ T cells were located within 25μm of such cells and confirming them as immediate neighbours.

**Fig. 4:**
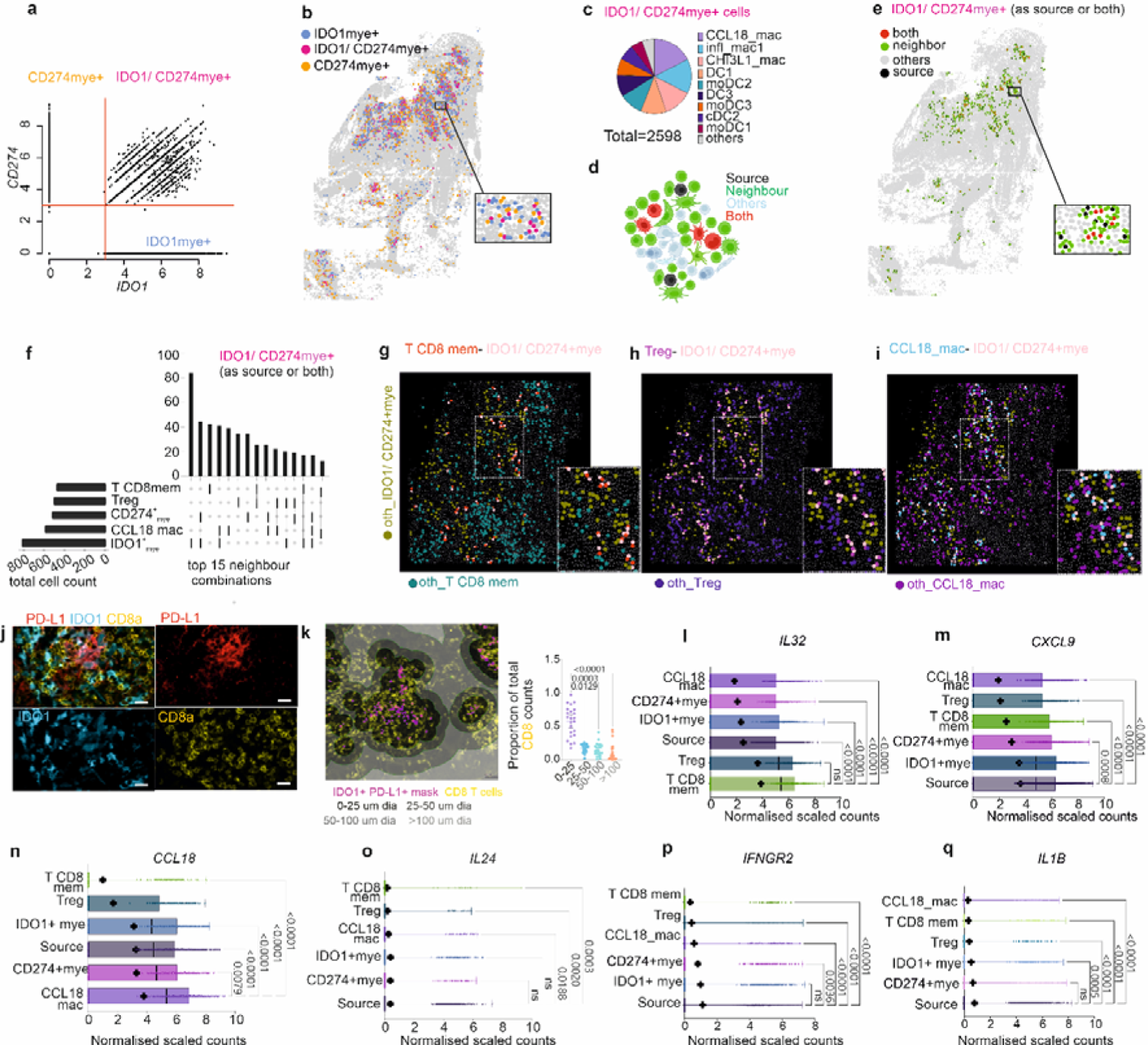
Phenotype of IDO1/ CD274+ myeloid cells and neighbours. **a**, Scatter plot of *CD274* and *IDO1* expression on myeloid cells in CosMx dataset. Thresholds at y=3 and x=3 were used to classify cells as single of double positive for IDO1 and CD274 (n=4 patients; P3-P6). **b,** Spatial plot for P3 coloured by class defined in (a). **c**, Pie chart of IDO1/ CD274mye^+^ cells by myeloid cell subset **d**, Cartoon representation of neighbourhood analysis (see M&M for details). **e**, Spatial map of IDO1^+^/ CD274mye^+^ as “source” or “both” shown with neighbours. **f**, UpsetR bar plot of cell types classed as neighbours of IDO1^+^/ CD274mye+ cells. Top 15 neighbour combinations are shown from 2418 source neighbour pairs. Connecting lines under the bar plot show neighbour combinations. Vertical bars represent total numbers of each combination while the horizontal plots indicate the total count of cell types as neighbours irrespective of the combination. **g-i**, Representative FOV showing spatial plots of IDO1^+^/ CD274mye^+^ cells (pink) interacting with CD8 mem (red dots, **g**), Treg (magenta, **h**) and CCL18_mac (blue in **i**). Other cells not in neighbour combinations are depicted with an “oth_” prefix below figure **j,** Immunohistochemistry showing expression of IDO1, PD-L1 and CD8A protein expression. **k**. Quantitative image analysis of (**j)** from n=23 patients. Left panel shows imaging masks in shades of grey at 25, 50 and 100 µm diameter from IDO1^+^ PD-L1^+^ cells. Right panel shows proportions of CD8^+^ cells by distance from IDO1^+^ PD-L1^+^ double positive cell. Friedman’s one-way test with Dunn’s adjustment for multiple comparisons. Adjusted p-values are given. **l-q,** Box and whisker plots showing minimum and maximum expression of *IL32* (**l**), *CXCL9* (**m**), *CCL18* (**n**), *IL24* (**o**), *IFNGR2* (p) and *IL1B* (q) in top 5 neighbours from **(f)**. One-way Kruskal Wallis test. Mean (+) and median (vertical line) are indicated inside bars. Dunn’s test corrected for multiple comparisons. Adjusted p-values are given.

We then examined the phenotypes of source and neighbouring cells with respect to cytokines and chemokines identified above as co-enriched in *CD274* and *IDO1* niches (**Fig. 3c**). *IL32* expression was significantly higher in CD8^+^ T memory cells and Tregs whereas *CXCL9* and *CCL18* were predominantly expressed by other myeloid cells including those that also expressed *CD274* and *IDO1* (**Fig. 4l-n**). In contrast, *IL24*, *IFNGR2* and *IL1B* were primarily expressed by *IDO1*^+^ and *CD274*^+^ and source myeloid cells (**Fig 4n-q**). Similar patterns were observed for neighbours of *CD274* and *IDO1* single positive cells (**Extended Data Fig. 6k,l**).

Hence, in Sri Lankan CL lesions, CD8^+^ memory T cells and Tregs (**Extended Data Fig. 6m**) represent the most common neighbours of *CD274*^+^ *IDO1*^+^ cells and are a major source of the *CD274*- and *IDO1*-inducing cytokine IL-32. Since both *IL32*β and *IL32*γ isoforms can induce either IDO1 or/and PD-L1 in macrophages ^58, 59, 60^, and IL-32γ expression has been reported in patients with *L. (V.) braziliensis* infection ^61^, we investigated which isoform was expressed in SL_CL lesions. Quantitative RT-PCR confirmed that *IL32*γ and *IL32*β were the most highly upregulated isoforms in lesion compared to healthy skin (**Extended Data Fig. 7**).

### *IL32* is a common spatial correlate of *CD274* and *IDO1* expression

Although differing mechanisms of immunopathology may occur across the leishmaniasis disease spectrum, IDO1 and PD-L1 have been consistently identified in transcriptomic studies ^32, 62, 63, 64, 65^. To assess whether the *CD274* and *IDO1* niche composition we identified in SL_CL patients was also seen in other forms of dermal leishmaniasis, we performed Visium analysis on biopsies from four Brazilian patients with *L. (V.) braziliensis* CL (**Fig. 5a, b, Extended Data Fig. 8a, b and Supplementary Table 2**) and two patients from India with *L. donovani* post kala-azar dermal leishmaniasis (PKDL) (**Fig. 5c,d**, **Extended Data** Fig. 8c, d and **Supplementary Table 3**). Histologically, we observed a diffuse cellular infiltrate in these samples (**Fig. 5b,d** and **Extended Data** Fig. 8b,d), and *CD274* and *IDO1* expression showed similar patterns within the papillary dermis (**Fig. 5e,f** and **Extended Data** Fig. 8e-g). Using the same analysis strategy (**Fig. 5g** and **Extended Data** Fig. 8h), we found that *IDO1*^hi^ *CD274*^hi^ spots were restricted mainly to the upper dermis (**Fig. 5h,i** and **Extended Data** Fig. 8i, j). *IL32* and *CXCL9* were strongly expressed in all IDO1/ CD274 spots in both datasets along with other cytokines (**Extended Data** Fig. 9a-f). Based on the top 50 correlates (top 10 shown in **Extended Data** Fig. 9g-j), we identified signatures associated with *IDO1* (*IL32, GBP5, GZMB, LYZ, SOD2, IFI30, FTH1, GBP1, SRGN, CTSS;* **Fig. 5j**) and *CD274 (IL32, LYZ, WARS, IF130, FTH1 GBP1, FCER1G*; **Fig. 5k**) expression across all three disease forms. A subset of 5 genes emerged as a core correlate across all three datasets, namely *IL32*, *LYZ* (Lysozyme), *GBP1* (Guanylate-Binding Protein 1), *IFI30* (lysosomal thiol reductase) and *FTH1* (Ferritin heavy chain 1).

**Fig. 5:**
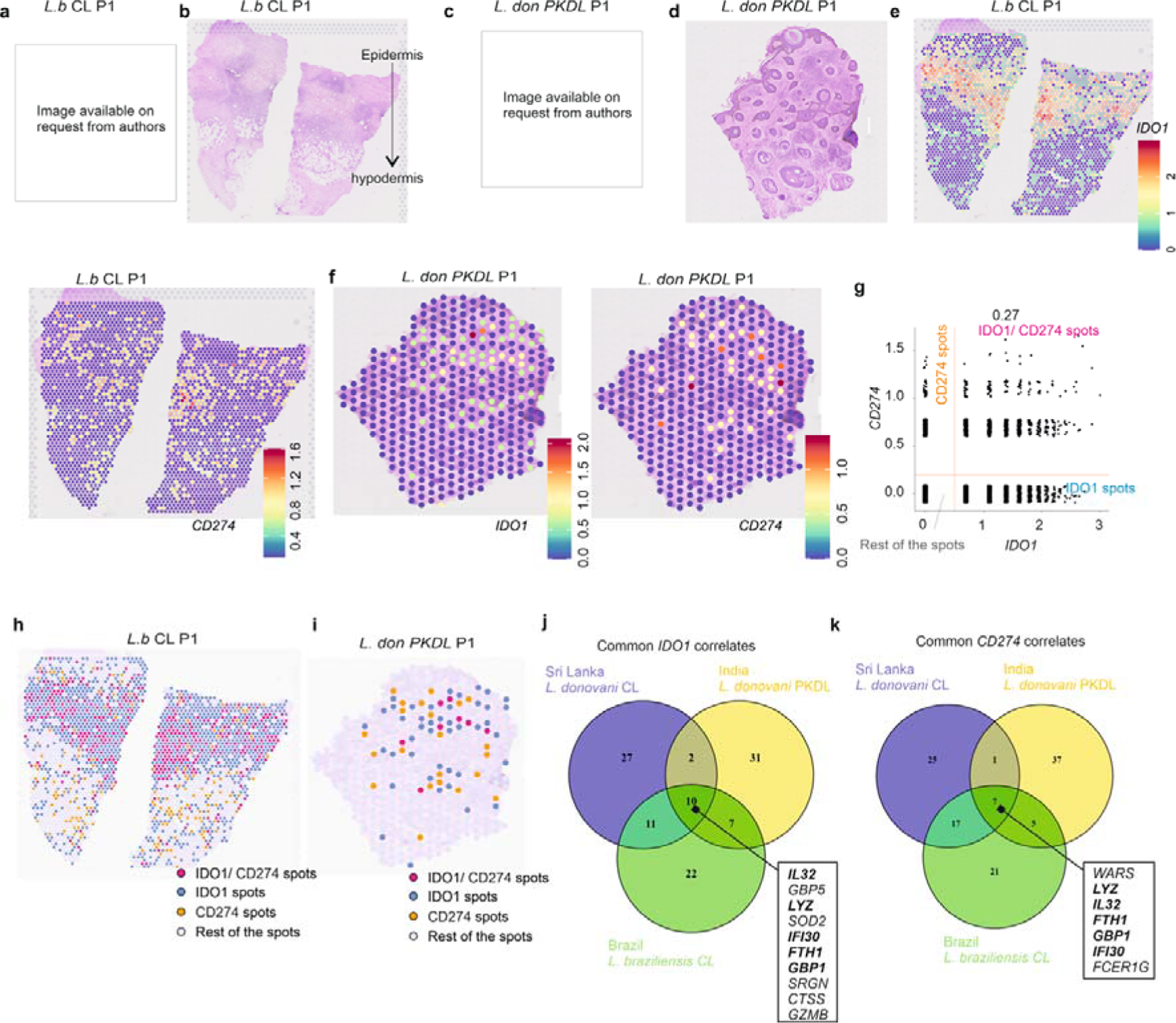
IDO1 and CD274 niches in different dermal variants of leishmaniasis. **a,** CL lesion caused by *L. braziliensis* in patient Lb_P1 from Brazil. **b**, H&E images of a 5μm section from (a) showing dense cell infiltration in the dermis. **C,** Polymorphic PKDL lesion caused by *L. donovani* in patient PKDL_P1 from India. **d,** H&E image of (c). **e-f,** Spatial plots showing the gene expression for *IDO1* and *CD274* on Lb_P1 (e) and PKDL_P1 (f). **g**, Scatter plot showing *CD274* and *IDO1* expression in all spots from *L. b* infected CL skin lesions (n=4). Thresholds at y=0.2 and x=0.5 used to classify spots. **h**, Spatial plot for *L.b* patients coloured by class as in (g). **i**, Spatial plot of IDO1 and PD-L1 classes for PKDL patients (n=2) derived from Extended Data Fig. 8h. **j-k**, Venn diagrams indicating overlap amongst top 50 genes per disease correlating with *IDO1* (**j**) and *CD274* (**k**).

### IL-32 is a predictive biomarker for rate of cure in Sri Lankan CL

As IL-32 has been previously shown to induce IDO1 and PD-L1 expression and the latter has been related to treatment outcome, we investigated the association between IL-32 expression and treatment outcome (**Fig. 6**). Quantitative analysis of IL-32 staining in the dermis (n=25 SL_CL patients; **Fig. 6a** and **Supplementary Table 1**) allowed stratification into two patient groups (IL-32^hi^ or IL-32^lo^) based on geomean number of IL-32^+^ cells / mm^2^ (**Fig. 6b**). IL-32^lo^ patients were significantly more likely to cure early after treatment compared to IL-32^hi^ patients (Log rank test, p=0.0025; **Fig. 6c**) with an estimated age- and sex-adjusted Cox model hazard ratio of 3.5 (95% CI, 1.19 – 10.3, p= 0.023; **Fig. 6d**). Given that our neighbourhood analysis above indicated a role for IL-32^+^ Treg cells in maintenance of the IDO / PD-L1 niche, we stained tissue sections for IL-32 in combination with a marker of Tregs (FoxP3; n=22 patients; **Fig. 6e**). Patients stratified based on FoxP3^+^ IL-32^+^ cell abundance (**Fig. 6f**) did not show a significant difference in cure rate (**Fig. 6g,h**). In contrast, when we conducted a similar analysis based on co-expression of IL-32 and CD8α (n=25 patients; **Fig. 6i-l**) we found that patients with low abundance of CD8^+^IL-32^+^ cells were significantly more likely to cure earlier (Log rank test, p=0051) with an age- and sex-adjusted Cox model hazard ratio of 2.78 (95% CI, 1.093-7.5, p=0.044; **Fig. 6k**). Collectively, these data strongly argue that IL-32^+^ CD8^+^ T cells are associated with the generation and / or maintenance of IDO1 and PD-L1 niches during CL and in such a way indirectly serve to restrain immune-dependent chemotherapy.

**Fig. 6.**
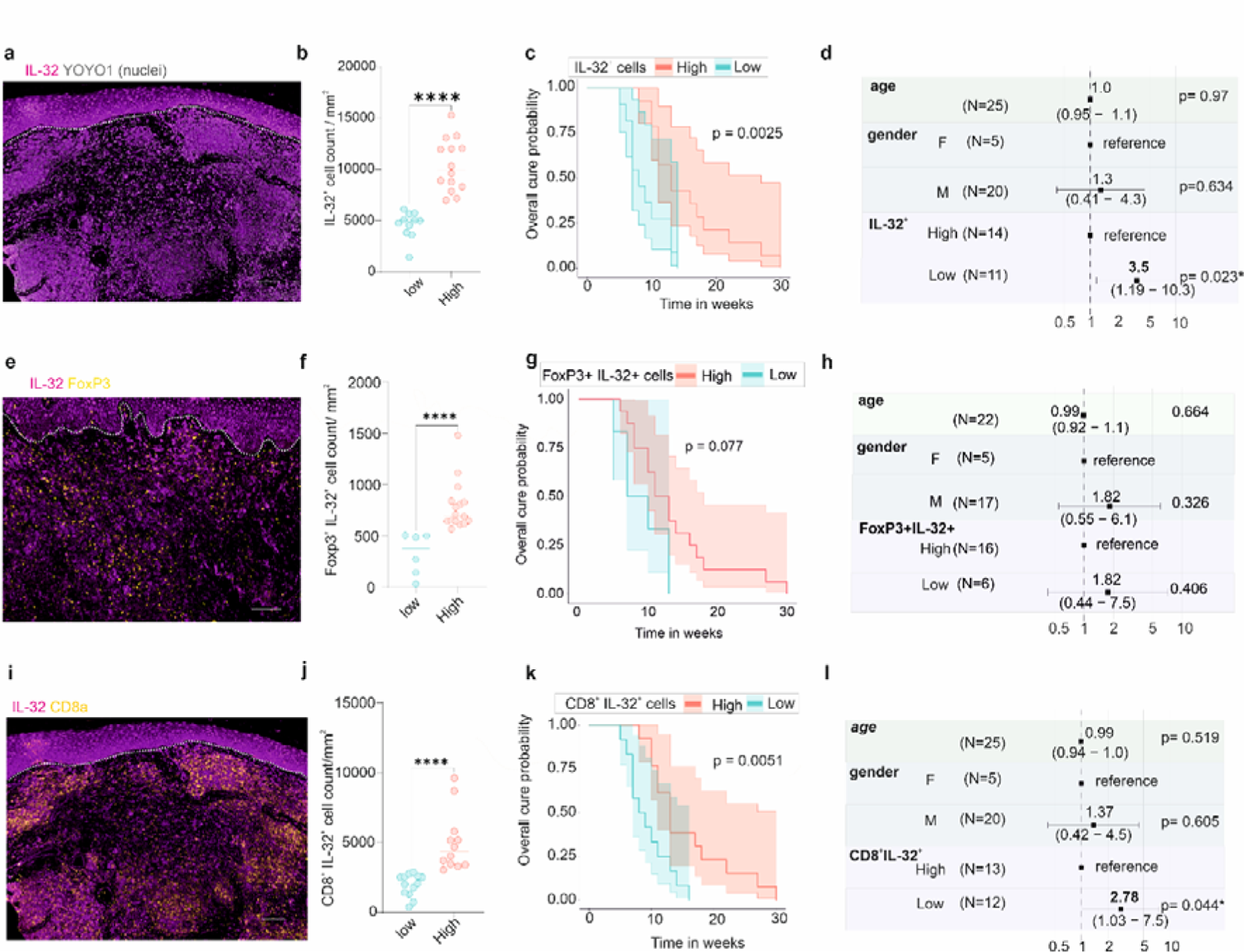
Lesion IL-32^+^ T cells are prognostic for rate of cure following treatment for CL in Sri Lanka. **a,** IL-32 protein expression in Sri Lankan CL. **b,** Stratification of 25 patients based on geomean density of dermal IL-32^+^ cells (low, n=11, High, n=14). Horizontal bars indicate median value. **** p-value <0.0001 (Two-tailed Mann Whitney test). **c,** Kaplan Meier plot showing treatment response rate for IL-32 Low and High groups. Shaded area indicates 95% CI. p-value for Log rank (Mantel-Cox) test is shown. **d,** Forest plot of Cox proportional hazard model for IL-32^+^ low vs. IL-32^+^ high (reference), adjusted for age and sex. Hazard ratio (HR; 95% CI) and p value for each covariate is shown. **e-h.** same as (a-d) for IL-32^+^ FoxP3^+^ cells (n=22 patients) **i-l,** same as (a-d) for IL-32^+^CD8α^+^ cells (n=25 patients). Scale bar in (a), (e), and (i) is 100 µm.s

## Discussion

Enrichment of cells expressing IDO1 and PD-L1 is a common feature of chronic inflammatory diseases ^66, 67, 68, 69, 70^ and cancer ^40, 71^, underscoring the role of myeloid-T cell interactions in pathological tissue microenvironments. High-throughput spatial profiling studies have revealed immunosuppressive niches in cancer tissue microenvironments (TMEs), comprising IDO1- and PD-L1-expressing suppressive macrophages, CD8^+^ T cells and regulatory T cells^33, 66, 72^. Two studies also shed light on the regulators of IDO1 and PD-L1 in the tumour microenvironment. One used multiplexed imaging to show IFNγ-dependent increases in CD4^+^/CD8^+^ T cells in *ex vivo* “responder” tumour explants ^66^. The second used limited protein/chemokine panels to characterize dysfunctional CD8^+^ T cells in melanoma-associated tertiary lymphoid structures ^73^. In our current study of CL lesions, we applied i) whole transcriptome-based spatial analysis to identify different cytokines and chemokines associated with IDO1 and PD-L1 microenvironments, ii) a targeted subcellular 1000-plex RNA panel to reveal cellular interactions at single cell resolution, and iii) validated our results using quantitative immunohistochemistry.

Our study provides new insights into the immunopathology of human CL. First, we identified myeloid cell-rich niches comprising mature dendritic cells (DC3) and *CCL18*^+^ M2-like macrophages (CCL18_mac) in the papillary dermis that highly express *CD274* (PD-L1) and *IDO1.* Second, neighbourhood analysis identified close interactions between *CD274*^+^/*IDO1*^+^ myeloid cells, and CD8^+^ memory T cells and Tregs as well as with *IDO1*^-^ *CD274*^-^ *CCL18*^+^ macrophages. Third, we identified a molecular signature associated with these niches (*CCL18, IL24, IL1B, TNFRSF6B, IFNGR2, CXCL9* and *IL32*) and their cellular source. Finally, using other geographically and clinically diverse forms of dermal leishmaniasis, we narrowed this signature to 5 common correlates of *IDO1* and *CD274* (*IL32, LYZ, GBP1, IFI30* and *FTH1*). While both GBP1^74^ and FTH1^75^ have garnered interest as therapeutic targets in cancer and inflammation, IL-32 emerged as a key factor produced by CD8^+^ T memory and Treg cells in our study. High abundance of IL-32^+^CD8^+^ T cells correlated with slower healing in Sri Lankan patients treated with sodium stibogluconate, identifying it as a prognostic biomarker for treatment response in this form of leishmaniasis. Hence, this study provides new insight into mechanisms supporting the generation of immunosuppressive niches in CL lesions and reveals cell interactions and gene signatures that may impact treatment response (as shown here) or disease natural history.

Myeloid cells play a central role in the immunobiology of leishmaniasis and have been explored extensively in mouse models, with studies ranging from seminal studies of migratory skin DCs ^76^ to the more recent identification of embryonic-derived CD206^hi^ macrophages ^77^ and inflammatory monocytes ^78^ as preferred host cells for *L. major*. Myeloid cell heterogeneity in the context of human leishmaniasis is much less well-understood and our analysis formally demonstrates the complexity of the myeloid cell response in terms of phenotypic heterogeneity and spatial organisation. We highlight two notable findings. First, we find a substantial proportion of moDCs and DCs distributed between the lesion core and T cell rich regions in the hypodermis. These DCs expresses *IDO1* and *CD274* supporting previous observations that DCs with regulatory phenotype are expanded in chronic inflammatory conditions and might contribute to immunosuppression ^38, 79^. Second, the lesion core contains abundant CCL18-expressing macrophages and these account for a significant fraction of all myeloid cells expressing *IDO1* and *CD274*. CCL18 produced by such macrophages may recruit naïve T cells ^80^ and Tregs ^48^ to further augment an inhibitory niche. In an analogous fashion, CCL18^+^ tumour associated macrophages (TAMs) promote immunosuppression in cancer ^81^. Furthermore, *IDO1*^+^ *CD274*^+^ *CCL18*^+^ macrophages express *IL1B*, *IL24*, and *IFNGR2.* Given previous studies that indicate a role for IFNs in the induction of PD-L1 and IDO1 in macrophages ^39, 42^ and for TNF in the induction of IL-24 and IL1B ^82^ these data are suggestive of macrophage activation by both IFNγ and TNF. This conclusion is further supported by co-expression of *TNF* and *IFNG* in IDO1/ CD274 spots in the lesion core, associated with T cell infiltration. Thus, in CL the regulation of IC expression appears distinct from that recently described in the IFNγ-depleted core of tuberculosis granulomas ^35^.

In the tumour microenvironment, macrophages also foster a supportive environment for malignant cell survival through coupled inhibition of apoptosis and compromised immunosurveillance ^83^. For example, soluble decoy receptor 3 (*TNFRSF6B;* DcR3) inhibits apoptosis by binding to LIGHT ^84^ or Fas-Fas ligand ^85^ complexes on T cells and, through its interaction with heparin sulphate proteoglycans, can also promote epigenetic reprogramming and macrophage polarisation to an M2 phenotype ^86^. We observed that DcR3 was also expressed in the IDO1 / PD-L1 niches found in CL lesions, suggesting that further exploration of the role of DcR3 in leishmaniasis is warranted.

A central finding of this study was the identification of IL-32 as a previously unknown core component of the IDO1 / PD-L1 niche in multiple forms of dermal leishmaniasis. IL-32 is a complex and still poorly understood cytokine that can be produced by multiple cell lineages (reviewed in ^87^). Studies of IL-32 are hampered by multiple isoforms, each with distinct biological effects and the lack of IL-32 in rodents ^88^. IL-32γ is the longest isoform with an intact signal peptide and is secreted. Both IL-32ß and IL-32γ are proinflammatory, with the beta isoform being the most abundant and the gamma isoform being the most biologically active ^89^. IL-32 has been mooted as a biomarker in a variety of skin conditions including atopic dermatitis and melanoma ^90^. It has been ascribed a pro-inflammatory and host protective role in tuberculosis and in models of colitis and arthritis based on the use of hu*IL32* transgenic mouse models. A role for IL-32 in leishmaniasis has also been previously suggested, based on similar transgenic mouse models and the enhanced expression of IL-32 in lesion biopsies from patients with *L. braziliensis* infection ^61, 88^. However, none of these published studies has addressed IL-32 expression in a spatially resolved manner. We confirmed high expression of both IL32γ and IL32ß in our patient biopsies, identified the cellular sources of IL-32 in the lesion, spatially mapped IL-32 and demonstrated that IL-32^+^ CD8^+^T cells represent a key molecular and cellular component of the IDO / PD-L1 niche. In keeping with other facets of IL-32 biology, our data imply a microenvironmentally-controlled immunoregulatory role for IL-32 in human CL.

Despite the differences in pathologies between SL_CL, *L. braziliensis* CL and PKDL, our analysis revealed a core gene signature (*IL32, LYZ, GBP1, IFI30* and *FTH1*) associated with *IDO1*^+^ and *CD274*^+^ expression. IL-32 expression is commonly associated with T cells and especially Tregs, though we demonstrate that this cytokine can also be expressed at low frequency in some myeloid cells in CL lesions. In contrast, LYZ (lysozyme), GBP1 (Guanylate-Binding Protein 1, an interferon inducible GTPase), *IFI30* (Interferon inducible lysosomal thiol reductase) and *FTH1* (Ferritin heavy chain 1) are all prototypic myeloid cell markers. This mixed cellular composition was reflected by deconvolution of cell abundances in our Visium data sets, wherein we observed a predominance of Treg, CD8^+^ T cells, macrophages and DCs. Further studies are required to determine whether other members of this core signature contribute to shaping the immunoregulatory environment in leishmaniasis.

In our previous study of SL_CL patients, we identified elevated expression of IDO1 and PD-L1 in pre-treatment biopsies and demonstrated that an early reduction in PD-L1 after treatment onset was predictive of rate of cure ^32^. Although providing a mechanistic basis for how ICs might restrict the efficacy of T cell-dependent chemotherapy, this study did not provide insights into the cellular or molecular mechanisms ultimately responsible for driving IC expression. The results presented here now allow us to propose a model for the regulation of ICs in CL, rooted in spatial interaction mapping and validated through multiple orthogonal approaches: 1) CCL18^+^ macrophages recruit IL-32^+^ CD8^+^ T cells and IL-32^+^ Tregs into a myeloid cell niche; 2) IL-32^+^ CD8^+^ T cells and IL-32^+^ Tregs induce expression of ICs in DCs and macrophages by paracrine signalling through the as-yet-uncharacterised IL-32 receptor, 3) IDO1^+^PD-L1^+^ myeloid cells respond to T cell derived cytokines (IFNγ, TNF) and secrete additional M2-polarising cytokines such as IL-24 and IL-1β, and 4) IDO1 and PD-L1 expressed by DCs and macrophages in the niche promote loss of effector T cell function, through tryptophan starvation and PD-1 signalling respectively (see Graphical Abstract).

The study has some limitations. Clinical sampling was limited on ethical grounds to two biopsies, so we have been unable to evaluate expression levels of IDO1 or PD-L1 at disease resolution. Although not statistically significant, patients with low abundance of IL-32^+^FoxP3^+^ Tregs also showed a trend towards faster healing, a result that deserves further exploration with larger sample sizes. Logistical constraints associated with studying patients in an endemic country setting impacted our ability to perform phenotypic and / or functional characterisation of cells isolated from lesion biopsies. Consequently, our cell deconvolution approaches may not reflect all cell phenotypes present in CL lesions due to the use of non-matched scRNA-seq data sets. As we were unable to isolate lesional mononuclear cells for in vitro analysis, we cannot formally address the question of T cell antigen specificity or extend our analysis of myeloid cells ex vivo. Finally, we have limited our current analysis to exploring cell-extrinsic regulation of IDO1 and PD-L1 expression by myeloid cells. Ongoing studies are exploring how the regulation of ICs on myeloid cells is regulated in a cell-intrinsic manner following intracellular parasitism by *Leishmania* (Dey et al, in preparation).

In summary, using spatially resolved high dimensional analysis of lesion biopsies, we have mapped the molecular and spatial niches associated with IDO1 and PD-L1 expressing myeloid cells in CL lesions and identified IL-32-expressing CD8^+^T cells as prognostic of rate of cure. Given the commonality of niche composition across multiple forms of dermal leishmaniasis, it is tempting to speculate that our observations may also contribute to local immunoregulation and treatment response in multiple forms of dermal leishmaniasis and that similar regulation of the IDO1/ PD-L1 niche may occur during other infectious and non-infectious diseases.

## Methods

### Samples: SL2 cohort, BR cohort and IN cohort. Study design

#### Sri Lanka

The study design for the Sri Lankan cohort was initially published as part of the validation cohort details in Dey et al, 2021 ^32^. Briefly, individuals who provided written informed consent and exhibited clinically highly suspected CL lesions were enrolled. On day 0, punch biopsies were collected and intralesional sodium stibogluconate (SSG) treatment was administered. Biopsies were processed into formalin-fixed and embedded in paraffin (FFPE) format and then shipped to the University of York (UoY) for subsequent immunological studies described here. These enrolled patients were then monitored for up to 6 months to assess complete clinical cure, and given weekly intralesional SSG injections. Of these, 24/25 patients had achieved complete clinical cure by 6 months, and 1/25 patients at 6.5 months. Out of the 25 patients in this cohort, 23/25 patient sections were included in the neighbourhood image analysis aimed at identifying neighbours of IDO1 and PD-L1 in stained sections; two were excluded due to insufficient material in the blocks. For including samples for 10x Visium analysis, these 23 patients were further screened for the presence of parasites by using an RNA probe against *Amastin* to detect parasites in 5u thick sections as described previously ^32^. 6/23 were *Amastin*^+^ and were included for Visium spatial analysis (P1-P6). Furthermore, out of the 6 samples included for Visium analysis, only four samples could be included in the CosMx analysis due to the availability of sufficient material in the blocks for follow up studies.

#### Brazil

Patient cohort from Brazil involved 20 patients that agreed to participate by signing the informed consent Form. Patients were diagnosed on the basis of positive polymerase chain reaction and/or indirect immunofluorescence and/or ELISA directed to *Leishmania*. Depending on the size and location of the lesion and the clinical condition of the patients, treatment was done with systemic pentavalent antimonials (Glucantime) for 20 days; or intralesional pentavalent antimonials (Glucantime) with one application per week for 4 weeks; or liposomal amphotericin B for 10 days. Biopsy samples of the lesion were collected on day 0 when the diagnosis was established and patients were followed up after treatment for 6 months to assess clinical cure. To understand IDO1 and CD274 microenvironments in healing and non-healing chronic skin *L.(V)b.* lesions, 20 patient samples were screened for double positivity against diagnostic PCR and ELISA (12/20) and were then stratified on the basis of visual treatment score at 3 months; healed if visual score=0 at 3 months and not healed if visual score >0 at 3 months. Of 12 samples, only 2 patients had healing score>0 at 3 months and were included in the study. 2 patients with visual score=0 at 3 months with comparable age, lesion size and lesion duration were then selected from the bigger cohort (12) to include a total of 4 samples that were processed through 10x Visium.

#### India

The initial diagnosis was based on clinical features suggestive of PKDL (presence of papules, nodules and/or hypopigmented macular lesions), a prior history of VL, rK-39 positivity, and/or if they resided in an area endemic for VL. In general, cases with hypopigmented patches were considered as macular PKDL, whereas cases with an assortment of papules, nodules, plaques, and/or macules were termed as polymorphic PKDL. Based on these criteria, individuals who were willing and provided written informed consent were enrolled for this study. The patients were examined by a clinician, clinical history recorded, and biological samples (blood and punch biopsy) collected. Subsequently, the diagnosis was confirmed by qPCR ^91^. 2 samples with typical histopathology of PKDL lesions and comparable RNA quality were chosen. These two patients were enrolled between November 2019 and August 2021. 5u thick sections were cut and processed through 10x Visium.

#### Lesion characteristics and diagnostic methods

For patients recruited in Sri Lanka and included in the study, lesions were dry nodular ulcerative (n=8), dry ulcers (n=6), dry ulcerated plaques (n=5), wet ulcers (n=2), papular satellite lesions (n=2), nodule with satellite lesions (n=1), ulcerated plaque with satellite lesions (n=1) with mean ulcerated area of 51.13 mm2 (± SD, 81.1) and mean area of induration of 1430.2 ( ± SD, 1966.9). In addition to clinical assessment, CL was diagnosed by slit skin smears (SSS) (18/23) and PCR (23/23).

Amongst Indian patients, the polymorphic case presented with lesions (papules and nodules) on the face, specifically on the forehead, nose and chin while the macular patient had a hypo pigmented patch on the back. In case of polymorphic cases, the biopsy was collected from a nodule, whereas in macular PKDL, it was from an area of hypopigmentation. In addition to clinical examination by a dermatologist, PKDL was also confirmed by qPCR. Both patients gave a previous history of VL, for which they had received sodium antimony gluconate For patients recruited in Brazil and included in the study, lesions were showed wet ulcers in the lower (3/4) or upper limbs (1/4) with mean ulcerated area of 2475mm2 (± SD, 2148). The diagnosis was confirmed by PCR specific for *Viannia* subgenus (4/4), indirect immunofluorescence (2/4) and ELISA (4/4).

#### Slit skin smears (SSS)

For Sri Lankan cohort only, tissue scrapings from a 3mm superficial nick from the active edge of the lesions were used to prepare smears on slides, stained with Giemsa and examined under oil immersion microscopy for the presence of amastigotes.

Parasite density was graded from 0 to 6+ according to WHO guidelines for VL^92^: 0—no parasites per 1000 high power fields (HPF: x 1000 magnification); 1+: 1–10 parasites per 1000 HPFs; 2+: 1–10 parasites per 100 HPFs; 3+: 1–10 parasites per 10 HPFs; 4+: 1–10 parasites per HPF; 5+: 10–100 parasites per HPF; and 6+: > 100 parasites per HPF. Amastigote density for each Sri Lankan sample is outlined in Supplementary Table 1.

#### Punch Biopsy

A 3mm diameter full thickness punch biopsy was taken from the edge or centre of the lesion under local anaesthesia, transported in formol saline and then fixed in paraffin blocks and used for H&E or FISH + IF and transcriptome analysis studies. Simultaneously an 2mm diameter punch biopsy was taken for PCR.

#### PCR

DNA extraction was performed from 2mm excision biopsy and was used as an input material for diagnostic PCR for *Leishmania* positivity as previously described ^32^.In Sri Lanka, LITSR/L5.8S specific primers were used that amplifies a 320 bp fragment of ITS1 region of Leishmania genus-specific DNA. In Brazil, kDNA specific primers were used that amplify 750 bp kinetoplastid DNA of *Leishmania* subgenus *Viannia* ^93^. In India, DNA extraction was performed using manufacturer’s instructions using QIAamp DNA mini kit, Qiagen, Hilden, Germany from a skin biopsy collected in phosphate-buffered saline (20 mM, pH 7.4) and eluted in 50 μL of DNA elution buffer. Real-time PCR was performed using specific primers for kinetoplast minicircle gene, using 1ul of DNA as input material and 400 nM of each primer. DNA isolated from a *Leishmania donovani* strain MHOM/IN/1983/AG83 served as the positive control while negative controls used were DNA from a healthy donor (no amplification), and a reaction mixture with water instead of template DNA (no-template control) ^91^.

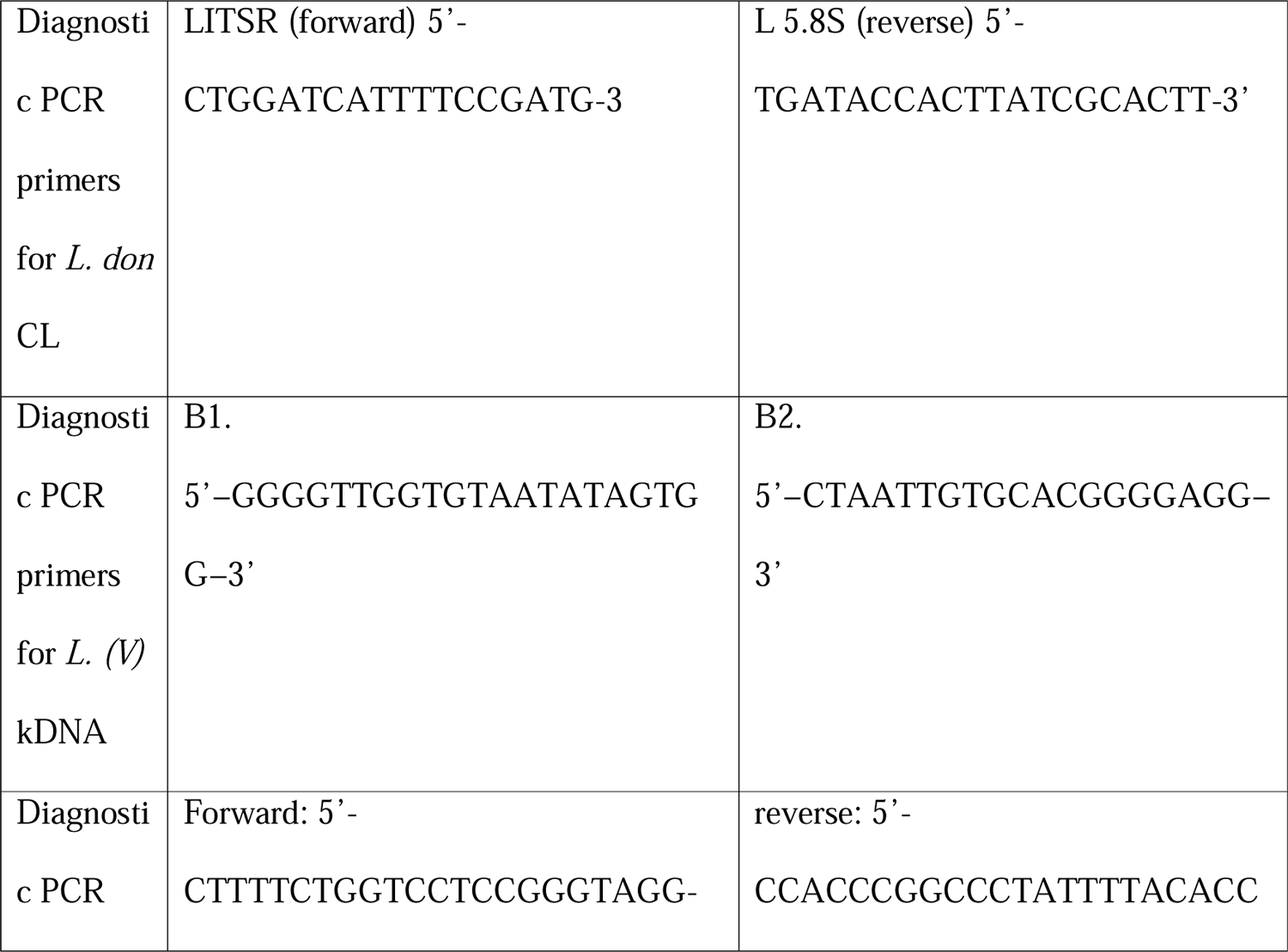
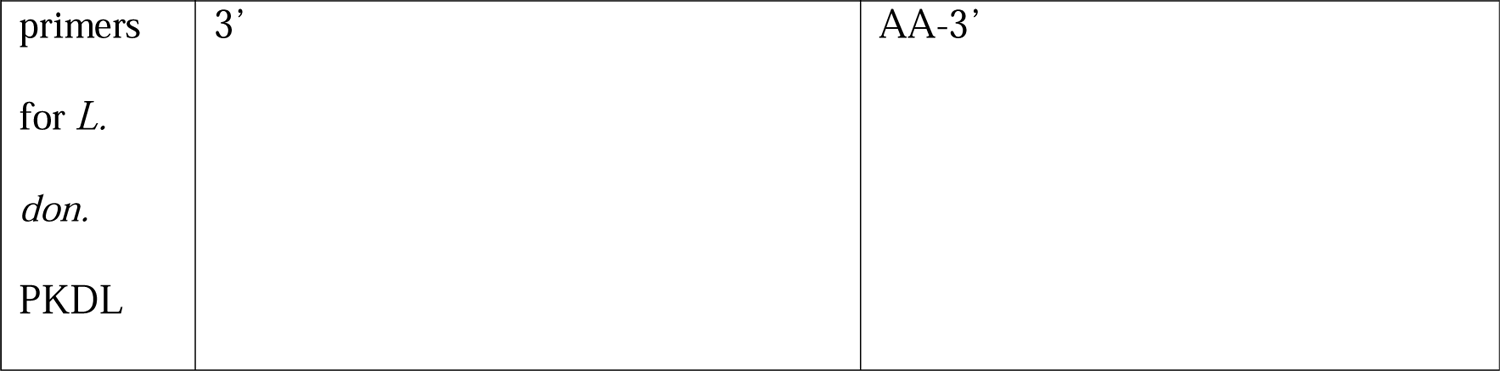

#### ELISA and Indirect immunofluorescence reaction

In Brazil, all sera were tested for immunoglobulin G (IgG) antibodies at 1:50 dilution in the ELISA using whole *L. major*-like parasite lysate (ELISA–*L. major*-like) as previously described ^94^. For both antigens, the cut-off point was determined using a receiver operating characteristic (ROC) curve. The reactivity index (RI) was calculated for each sample by dividing the sample absorbance value by the cut-off. Samples were considered positive if the RI value was ≥ 1. The spectrophotometric reading of each well was performed at 450 nm using a Multiskan GO instrument (Thermo Scientific, Finland). The IIF test was performed on the slides, containing the suspension of fixed promastigotes of *Leishmania* (*Leishmania major*-like promastigotes/MHOM/BR/71/49) as previously described ^95^. Then the slide was observed in the fluorescence microscope, with the objective of 250 ×.

### Sectioning

FFPE blocks were placed on cool plate prior to sectioning and 5μm sections were cut and serial sections were placed onto Superfrost glass slides for CosMx or H&E or 10x Visium onto the capture area making sure that all layers of the skin were represented in the cut section.

### Low-resolution spatial transcriptomic mapping of CL lesions using 10x Visium

Visium gene expression slides were processed using the Visium Spatial gene expression reagent kits for FFPE as per recommended protocols. In brief, slides were deparaffinised in Xylene twice for 10 minutes each and then stained for hematoxylin and eosin. Stained slides were coverslipped and imaged on an Axioscan slide scanner. Post imaging cover slips were removed, slides were placed in a Visium cassette and were de-crosslinked. Human probes were added overnight after which they were extended and released. Library preparation was done based on manufacturer’s instructions and sequenced using a NovaSeq 6000. Raw FASTQ files were aligned to the GRCh38 genome (https://www.ncbi.nlm.nih.gov/datasets/genome/GCF_000001405.40/) and the Spaceranger_1.3.0 (10x) count was used to generate UMI counts per spatial barcode. Raw counts were normalised and analysed further. Code used for analysis and figures is described under “Code Availability”.

### Integration and clustering of 10x Visium data

We used Seurat (Seurat_4.3.0) package in R 4.2.3 to analyse spot matrices. Normalisation across spots was performed using SCTransform(). We used FindIntegrationAnchors() function to find anchor spots across samples and then used IntegrateData() to integrate all samples based on 3000 features as identified upon data integration. 30 Principal Components derived using 3000 features in the integrated assay were used as an input for clustering Visium spots with a resolution of 0.5 using FindClusters() and UMAP was used to visualise clustering in two-dimensional space. Differential gene expression (DE)between clusters was calculated first by running PrepSCTFindMarkers() to normalise for median genes as a proxy for sequencing depth. FindAllMarkers() was then used to find DE genes using a minimum setting for log2fold change as 0.25 with genes that were expressed in a minimum of 25% spots (min.pct=0.25). P-value of DE genes was based on a two-tailed Wilcoxon Rank Sum test. List of DE markers from each cluster was then fed into CellMesh ^96^ ( https://uncurl.cs.washington.edu/db_query) using probabilistic model to identify probable cell types expressed in cluster spots. Clusters were then annotated based on most abundant cell type expressed and then plotted onto each tissue image using SpatialDimPlot function from Seurat and required DE genes were visualised using EnhancedVolcano package in R. Spatial maps of *IDO1* and *CD274* (PD-L1) were generated using SpatialPlot() function. For comparison across My1, My2 and My3, matrices were extracted from Seurat and plotted in Graphpad for violin plots.

### Deconvolution of Visium spots to identify immune cell abundance

We utilized Cell2location ^52^ to deconvolve cell type composition per 55μm Visium spatial spots. The hyperparameters used were N_cells_per_location=30 and detection_alpha=20, using the dataset Reynolds *et al* ^38^ (accession number: E-MTAB-8142; www.ebi.ac.uk/arrayexpress/experiments/E-MTAB-8142.) as a skin single-cell reference dataset. First, we estimated reference cell type signatures by training a negative binomial regression model for 1,000 epochs retaining the cell-type annotations from *Reynolds et al*. We then used the reference signature model to deconvolute the spatial data using the aforementioned hyperparameters, training the Cell2location model for 30,000 epochs. We inferred cell abundance per spatial spot using the 5% quantile values of the posterior distribution, where the model has high confidence as per the tool’s manual. Predicted abundances were used to infer compositions in My1, My2, My3, IDO1, CD274 and IDO1/ CD274 spots.

### Identifying molecular networks associated with CD274 and IDO1 expression

To look for correlation between *IDO1* and *CD274* across all spots, all spots covered under the tissue from Sri Lankan patients (n=6) were visualised using FeatureScatter() function in Seurat. Thresholds on the scatterplot were put below or equal to median of total expression of *IDO1* (1.2) and *CD274* (0.6). All spots were then annotated as IDO1 spots (*IDO1* > 1.1 & *CD274* < 0.5) or CD274 spots (*IDO1* < 1.1 & *CD274* > 0.5) or IDO1/ CD274 spots (*IDO1* > 1.1 & *CD274* > 0.5) or rest of the spots (*IDO1* < 1.1 & *CD274*< 0.5). DE genes were then graphed as line plot in GraphPad 9.3.1 whereas the rest were plotted as dot plots using DotPlot function in Seurat in R. A similar strategy was followed for patients from Brazil (n=4) presenting with CL lesions and patients from India (n=2) presenting with PKDL lesions.

We performed a pairwise correlation analysis of all genes in the dataset with *IDO1* and *CD274* using the cor.test() function from the R stats package (version 4.1.1), employing Spearman’s ranked correlation method. A significance threshold of 0.05 was applied to identify significant correlations. Subsequently, the top 50 correlated genes from each dataset, which included Visium gene expression data from 6 Sri Lankan CL patients, 4 CL patients from Brazil, and 2 PKDL patients from India, were extracted. These gene sets were then subjected to comparison using Venny 2.0 (https://bioinfogp.cnb.csic.es/tools/venny/index2.0.2.html) to identify common correlates of *IDO1* and *CD274* across these three datasets.

### Single cell resolution spatial transcriptomic mapping of CL lesions using CosMx molecular imager

For CosMx, a single slide containing sections from skin lesions of 4 patients from SL2 patients were sent to Nanostring Translational Services lab, Seattle under Technology Access Program (TAP). CosMx Human Universal Cell Characterization RNA Panel (1000-plex) was used to analyse gene expression at subcellular level. Morphology of the tissue was visualised by staining for B2M/CD298, PanCK, OPB (for parasite), CD3 antibodies and DAPI post hybridization of probes. The methods for sample processing and cell segmentation are described in CosMx methods study ^53^. A total of 20 Fields of View (FOVs) were created across 4 patient samples. Expression matrix of transcripts along with their x, y and z coordinates, cell ID, metadata, FOV positions were loaded onto Giotto package to create an integrated giotto object in R ^56^ and then filtered based on minimum expression threshold of 1;> 5 features/cell and > 5 cells expressing one feature. Expression matrix was normalised using standard method of data normalisation for total library size followed by scaling for a factor of 6000, log normalisation and z scoring of data by genes and/or cells. Expression values were then regressed for total sum of feature expression and number of features expressed per cell to account for batch to batch variation.

### Clustering and cell type annotation of CosMx data

UMAP dimension reduction algorithm was used for individual cell data presentation based on CosMx SMI gene expression profiles. For a further QC, total count numbers and slide ID were projected onto UMAP space to assess data quality and no total count-based clustering bias was found. Coarse cell typing was done using nb_clust, a semi-supervised cell clustering (negative binomial) algorithm developed by NanoString team resulting in detection of 16 immune cell types using single cell data sets. 6 unsupervised cell clusters (KC1, KC2, KC3, plasmablast2, fib2 and mac2) were also identified and were labelled on the most enriched cell type based on CellMesh reference database ^96^. DE between clusters was done using findMarkers_one_vs_all from Giotto package using Gini coefficient Followed by plotMetaDataHeatmap from Giotto package to plot heatmap of gene expression. Cell ID and expression matrix of all cells were then imported into Seurat package and subclustered to identify further subtypes. Myeloid cells (cluster IDs: mac, mac2, mDC, neutrophil, pDC, monocyte, mast) were subclustered into13 subtypes that were annotated on gene expression and projected onto UMAP space. 12% of total cells that showed abundance of border transcripts from nearby cells due to faulty assignment of transcripts were subsetted out. ImageDimplot from Seurat and SpatPlot from Giotto were used to map cell types onto space.

### Neighbourhood analysis of *CD274* and *IDO1* expressing cells

*IDO1* and *CD274* values from all myeloid cells were correlated and thresholds were placed that showed clear assignment of all myeloid cells into IDO1mye+ (*IDO1* > 3 and *CD274* <3), CD274mye+ (*IDO1* < 3 and *CD274* >3), IDO1^+^/CD274mye+ (*IDO1* > 3 and *CD274* >3) and were annotated as such. SpatPlot2D from Giotto package was used to map cell types onto space. To create a network that revealed spatial relationships and interactions among individual cells, we first used plotStatDelaunayNetwork function from Giotto to estimate distance between each cell and its neighbours by calculating pairwise distances between cells and identifying statistically significant connections. We estimated that each cell had 4-6 nearest neighbours. We then used createSpatialNetwork from Giotto using k =4 to limit the number of neighbours to 4.

To investigate cellular interactions within the spatial network, we first created an interaction map for IDO1mye^+^, CD274mye^+^, IDO1^+^/ CD274mye^+^ with 4 immediate neighbour cells using annotateSpatialNetwork function from Giotto. To create spatial plots illustrating these interactions, we first used findNetworkNeighbors function to identify these neighbours.

Spatial plots were then generated using SpatPlot2D function to visualize the cellular interactions. Cells were color-coded based on their classification as “source” (specified cell; IDO1mye^+^ or CD274mye^+^ or IDO1^+^/CD274mye^+^) or “neighbour” (amongst 4-6 immediate neighbours of specified cell) or “both” (a neighbour and a source cell) or “others” (not a neighbour to specified cell nor a source cell). Cell IDs and cell type annotation of these cells were then imported into UpsetR shiny app ^57^ (https://gehlenborglab.shinyapps.io/upsetr/) to estimate the most abundant neighbour combination to IDO1mye^+^ or CD274mye^+^ or IDO1^+^/CD274mye^+^ cells and into a worksheet to estimate composition of these populations and graphed in GraphPad. For additional validation, neighbour combinations were also checked in an excel sheet to tally neighbour combination numbers. To look at spatial maps of specific interactions, an additional metadata column with information about interacting and non-interacting cell types of the selected cell-cell interaction was created and visualised using spatPlot from Giotto.

To look at cytokine and chemokine expression patterns in source and neighbouring cells, cell IDs of IDO1mye+ or CD274mye+ or IDO1^+^/CD274mye+ source cells and top 5 neighbours (CCL18_mac, Treg, IDO1mye+, CD274mye+, T CD8 memory) were subsetted and violin plots, heatmaps (using Giotto in R) or bar charts (in GraphPad Prism 10) were plotted to look at cytokine expression in these cells.

### Immunostaining

FFPE sections were obtained from 23 or 25 Sri Lankan patients, with 23 FFPE patient lesion blocks used for the IDO1, PD-L1, and CD8 staining set due to material constraints. For FoxP3 staining set, only 22 sections were analysed due to sample detachment during processing. The staining protocol involved following steps: heat fixation at 60°C to ensure sample adherence, followed by two rounds of 5-minute deparaffinization in histo clear/xylene at room temperature (RT). Subsequent steps included equilibration in 95% ethanol for 3 minutes, 70% ethanol for another 3 minutes, and hydration in distilled water for 3 minutes. Antigen retrieval was achieved through a 15-minute high-pressure treatment with citrate buffer (pH 6.0), followed by a 25-minute standing period at RT. After two 5-minute washes with wash buffer (PBS + 0.5% BSA w/v), tissue sections were delineated with a wax pen. The procedure continued with blocking (PBS + 0.1% Triton X-100 v/v + 5% Normal donkey serum v/v + 5% Normal goat serum v/v + 1% BSA w/v) for 30 minutes, overnight primary antibody incubation with IDO1 (1:200) and PD-L1 (1:500) at 4°C in the fridge, and three 5-minute washes the next day. Secondary antibody staining for 30-45 minutes at RT employed CF750 Donkey Anti-Rabbit IgG CF™ 750 and Goat anti-Mouse Dy650 at a 1:500 concentration. Sections were then incubated with CD8-AF594 for 2h at room temperature.

After three more buffer washes, YOYO-1 at 0.2uM concentration was added for 30 minutes in wash buffer. Three final wash buffer washes were performed before applying Prolong Gold mounting media and coverslips.

For the IL-32 CD8 set and IL-32 FoxP3, the same protocol was followed with minor adjustments. Antigen retrieval utilized TE buffer (10mM Tris Base, 1mM EDTA, 0.05% Tween 20, pH 9.0), while pre-primary blocking consisted of PBS + 0.1% Triton X-100 v/v + 5% Normal goat serum v/v + 1% BSA w/v. The slides were incubated with IL-32 (1:400) and CD8a or FoxP3 overnight at 4°C and were developed with goat anti-rabbit Dylight 650 and Goat anti-mouse AF555 F(ab)2 at a 1:500 concentration, respectively. Notably, no second blocking step was carried out for IL-32 CD8/ FoxP3 set. Detailed antibody information, including catalogue numbers and dilutions, can be found in the reporting summary.

### Image acquisition and analysis

Whole tissue section mages were acquired using Zeiss AxioScan.Z1 slide scanner at 20x magnification. Identical exposure times and threshold settings were used for each channel on all sections of similar experiments. Whole section images of IDO1, PD-L1, CD8/ FoxP3 and IL-32 was were quantitated using StrataQuest Analysis Software (TissueGnostics, v 7.0.1.178). We first segmented nuclei based on YOYO1 staining and then created cell polygons over grey images of CD8 (outside), PD-L1(outside and inside) and IDO1 (outside and inside) using nuclei mask. A cut off was applied on IDO1/PDL-1 mean intensity based on visual inspection of the tissue to delineate IDO1/ PD-L1 ^+^/^-^ cells. We then created coded images for each of the markers and used proximity maps function in Strataquest 7.178 (https://tissuegnostics.com/products/contextual-image-analysis/strataquest) to generate distance maps at 25, 50 and 100 µm from IDO1, PD-L1 or IDO1 PD-L1 double positive polygons and then detected CD8 masks within these proximity areas For quantification of IL-32 and CD8/ FoxP3, nuclear segmentation was based on YOYO1 and expanded outside of nuclei for 10 µm for IL-32 and outside for 1µm for CD8 (also a ring mask with interior and exterior radius of −0.20 and +0.20 um). As FoxP3 staining was nuclear, cell polygons were created on FoxP3 staining without any expansion on nuclei mask. Cut-offs were also applied to nuclei, IL-32 and CD8/ FoxP3 mean intensity detection to ensure no false positives were included. The same cut-off was applied for all samples. Scattergram was generated with IL-32 and CD8/ FoxP3 mean intensities and gated on positive IL-32 and CD8/ FoxP3 cells. Upper right quadrant of these scattergrams gave data for IL-32^+^ CD8^+^/ IL-32^+^ FoxP3^+^ cells.

### Multivariate Cox proportional hazard model

For correlation of IL-32 with cure rates, total IL-32^+,^ CD8^+^ IL-32^+^ and FoxP3^+^ IL-32^+^ cells per mm2 of tissue section per patient (n=22-25) was computed from image analysis. Patients were then stratified on geomean of total IL-32 (6969.37), CD8^+^ IL-32^+^ (2800.9), FoxP3^+^ IL-32^+^ (544.0) expression into IL-32 high (>6969.37; n=14) or low (<6969.37; n=11) and CD8^+^IL-32^+^ high (>2800.9; n=13) or low (<2800.9; n=12) and FoxP3^+^IL-32^+^ high (>544.0; n=16) or low (<544; n=6). These values were then imported in R and Kaplan Meier survival (cure) curves were generated for IL-32or CD8^+^IL-32^+^ or FoxP3^+^ IL-32^+^ expression groups using the survfit function, and survival differences were assessed with the survdiff function from survminer, survival, ggplot2 packages. We then constructed multivariate hazard models adjusted for participant age and sex, employing the survival and survminer packages. The statistical scores were derived from Wald’s statistic value, followed by estimation of hazard ratios, which were visualized on forest plots.

### RNA isolation and qPCR

The total RNA was extracted from formalin-fixed paraffin-embedded (FFPE) patient samples using RNeasy FFPE Kit (Qiagen) as per manufacturer’s protocol. Briefly, 2 x 10µm sections were cut from each block and put into deparaffinization buffer (Qiagen). Sample lysis was done using Proteinase K digestion for 15 minutes. After lysis, samples were incubated at 80°C for 15 minutes followed by 15 minutes of DNase treatment. Finally, concentrated RNA was purified using RNeasy MinElute spin columns, and eluted in a volume of 14–30 µl.

Purity and concentration of extracted RNAs were checked and quantified by reading at 260 and 280 nm in a NanoDrop spectrophotometer (Thermo Fisher). Next, 50ng total RNA was reverse transcribed using Superscript IV first strand cDNA Synthesis Kit (Invitrogen) and random hexamers (Invitrogen) according to manufacturer’s protocol. qRT-PCR was performed with Fast SYBR Green Master Mix (ThermoFisher) and primers using a StepOnePlus Real[Time PCR System (ThermoFisher). cDNA extracted from LPS (100 ng/ul for 4 hours) stimulated PMA (100 ng/ul) differentiated THP-1 cells line was used as a positive control to optimise primers^97^. All primers were optimised by serially diluting LPS stimulated THP-1 cDNA. Relative transcript levels were determined using the ΔΔCt method using GAPDH/ ACTB as a housekeeping gene and healthy skin. All reactions were performed in duplicates. Primer list used in the study is given in the table below.

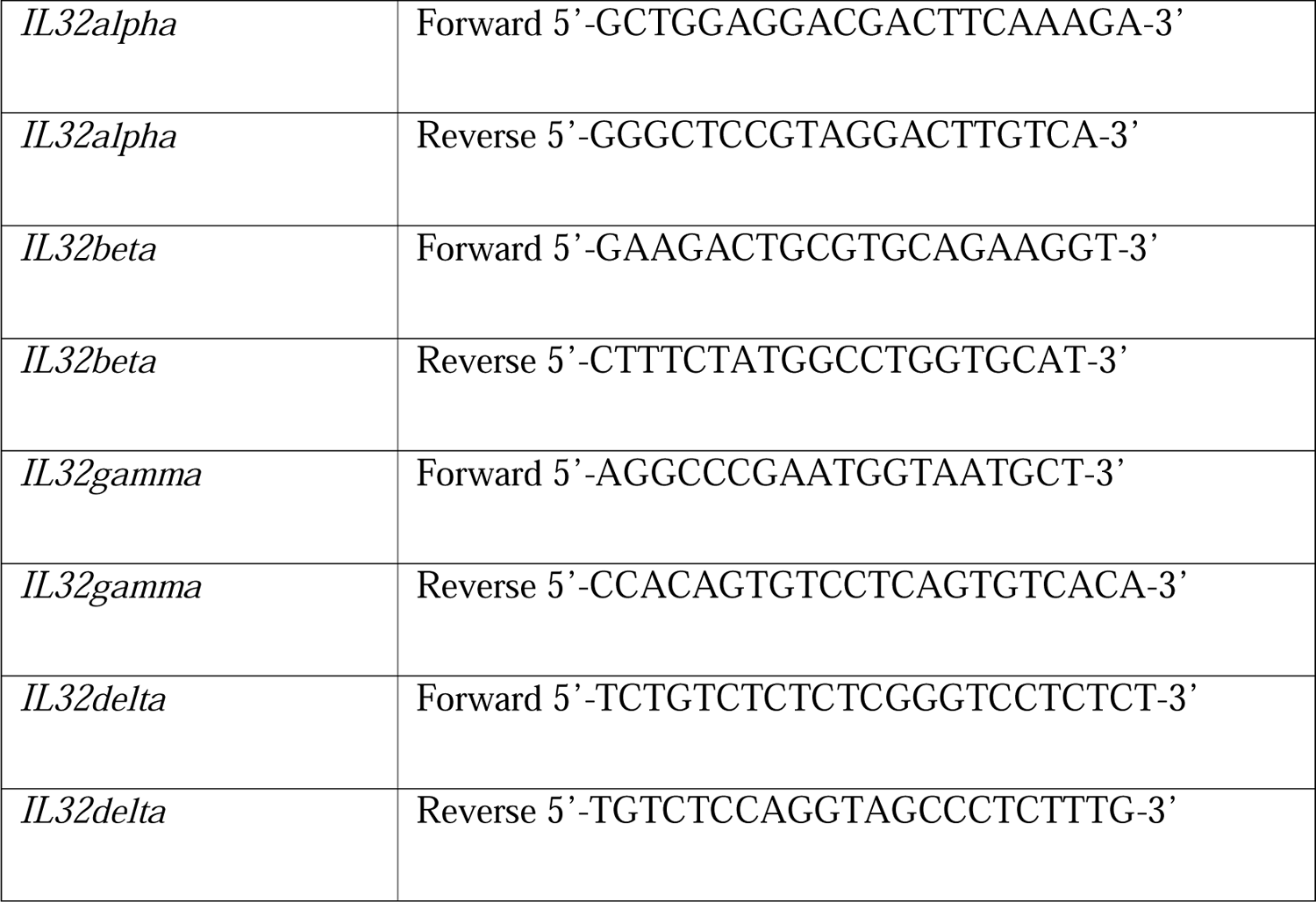

## Data Availability

The sequencing data reported in this paper have been deposited in the GEO database under the accession code GEO: TBC. The processed sequencing data is available as Rds files on Zenodo: https://doi.org/10.5281/zenodo.10402126. Additional data (**Supplementary Table 4**) related to the figures is also available on figshare (DOI: https://doi.org/10.6084/m9.figshare.24803511).

## Acknowledgements

The authors thank all patients and their families who took part in this study. We also thank all members of the Kaye laboratory at Hull York Medical School, University of York for their useful comments and suggestions on this project, Robert Nica and Simina Laslau at the technical department at Tissue Gnostics for help with Strataquest, TAP team at Nanostring, Seattle specially Emily Killingbeck, Youngmi Kim and Claire Williams for initial support with CosMx and the Biosciences Technology Facility at University of York for support throughout the project. Biorender.com was used to generate the graphical abstract.

## Author contributions

N.S.D., S.D. and P.M.K. conceived the study; N.S.D. and S.D. did the analysis; N.S.D., S.D., and N.B. conducted experiments; N.S.D and P.M.K wrote the manuscript; P.M.K, S.R., M.C., and H.G. secured funding; all other co-authors provided materials or clinical assessments for the study and reviewed the manuscript.

## Code availability

Code used for analysis (including that for generating Figs from Visium seq data and CosMx imaging data) in this study is available at https://github.com/NidhiSDey/leish-ME

## Ethics statements

### Institutional review board statement

Written informed consent including for lesion photographs was obtained from participants under a protocol for the project titled “Towards a global research network for the molecular pathological stratification of leishmaniasis” approved by Ethical Review Committee of the Faculty of Medical Sciences, University of Jayewardenepura (ref: 780/13 & 52/17), Ethics Committee of the Faculdade de Medicina, Universidade de Sao Paulo-CAAE 39964520.8.0000.0068, Institutional Ethics Committee of School of Tropical Medicine, Kolkata and Institute of Post Graduate Medical Education and Research, Kolkata (IPGME&R/IEC/2019/208) and the Biology Ethics Committee (ref: PK201805), Department of Biology, University of York. This study was conducted according to the Declaration of Helsinki (2013).

### Inclusion and Ethics statement

We affirm that our research upholds the highest ethical standards and contributes to the advancement of immunological science in an inclusive and responsible manner.

## Extended Figures

**Extended Data Fig 1:**
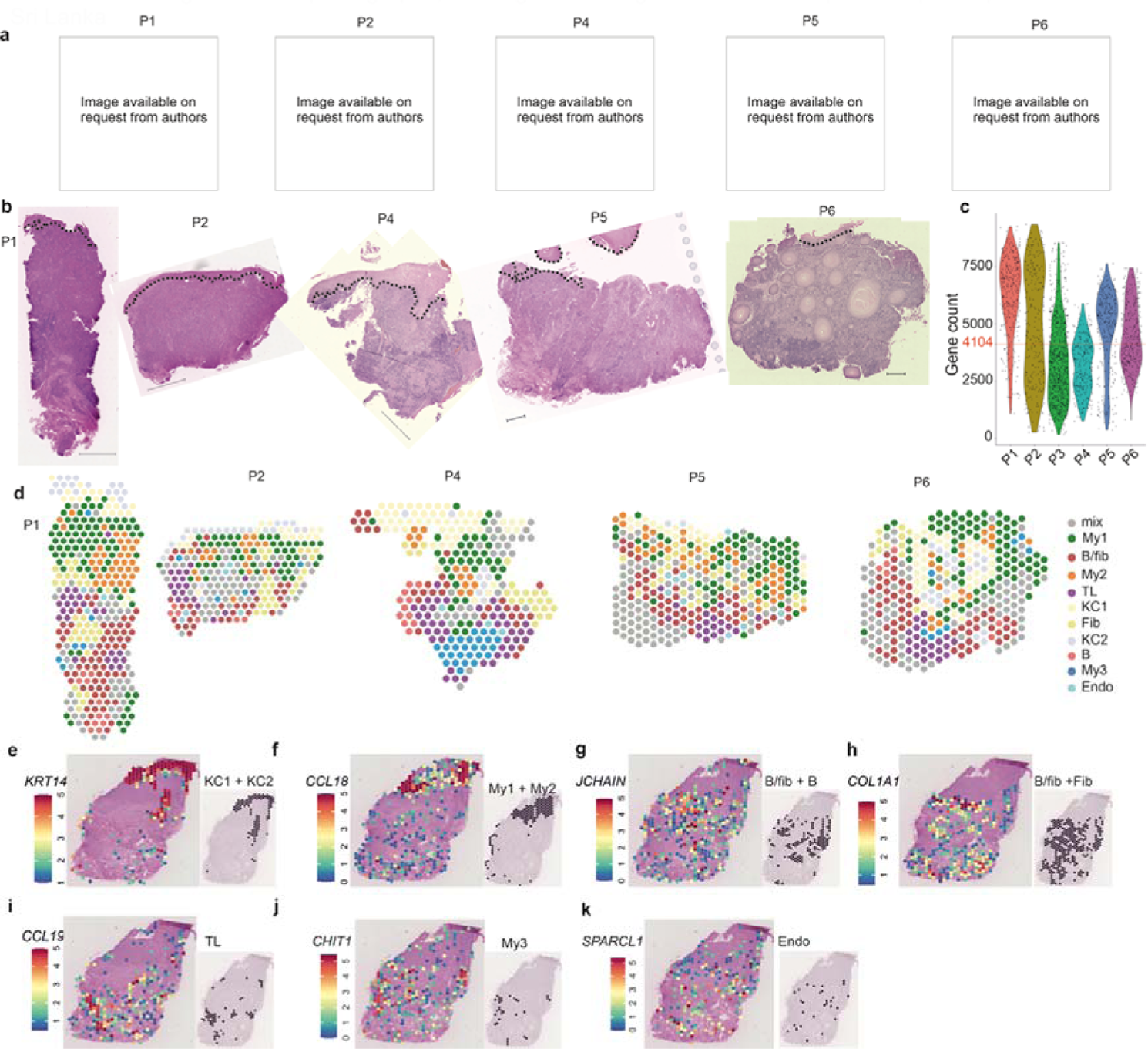
Spatial transcriptomics identifies spatial domains in *Leishmania donovani* infected CL skin. **a**, Photographs of patients P1, P2, P4, P5 and P6. **b**, H&E images of patients described in (**a**) showing dense cellular infiltrate. **c**, Violin plot showing overall gene counts across all patients. Red line shows median gene count across all samples (n=6) **d**, Spatial maps for patients described in (**a)** coloured by clusters identified in Fig.1f. **e-k,** Spatial feature plot for P3 overlaid on H&E showing normalised gene expression of top genes expressed in each cluster: *KRT14*, *CCL18*, *JCHAIN*, *COL1A1*, *CCL19*, *CHIT1* and *SPARCL1* (left panels) and spatial maps of respective clusters (right panels).

**Extended Data Fig. 2:**
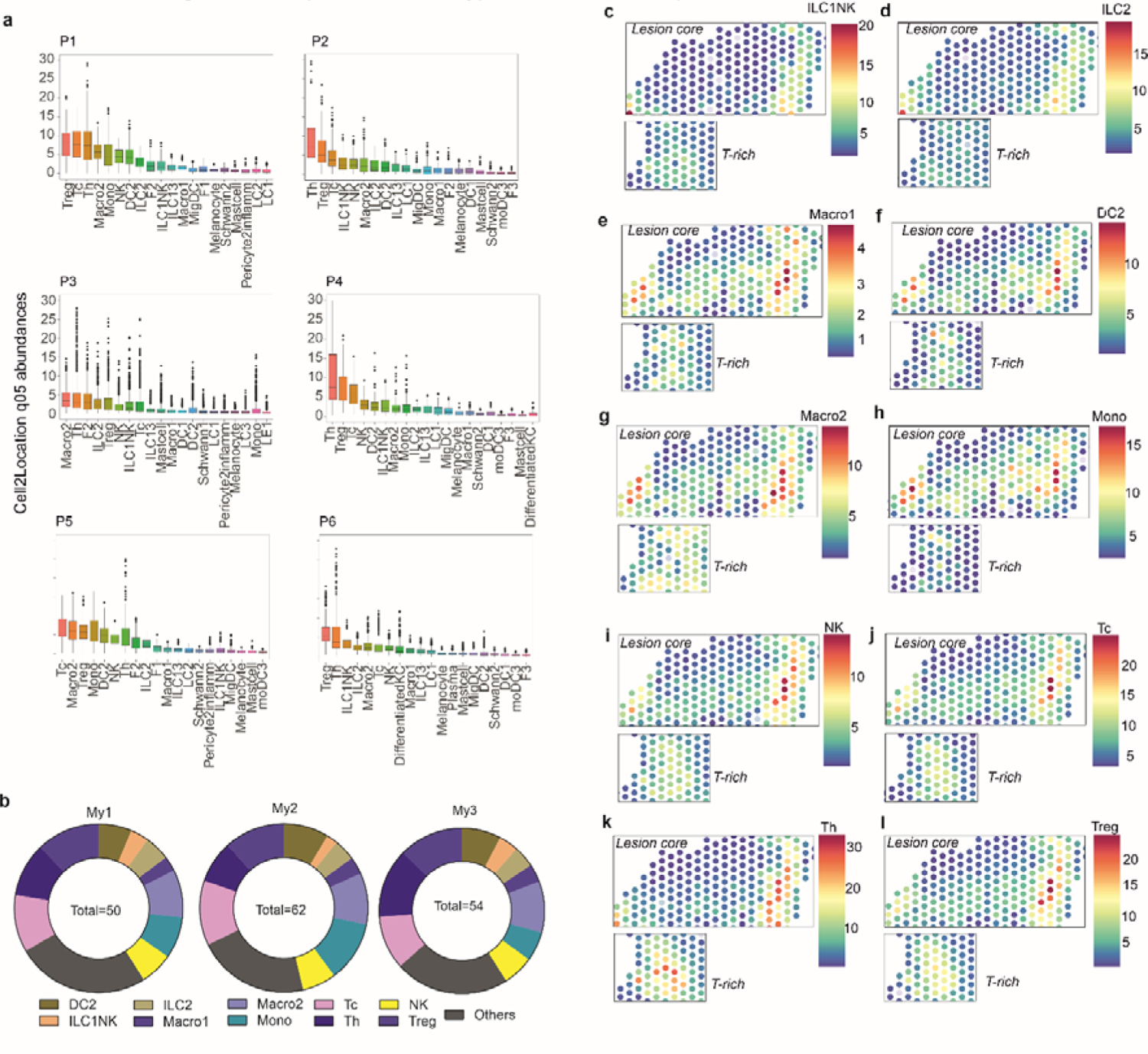
Deconvolution of cell types in 10x Visium spots. **a,** Bar plot showing 5^th^ quantile predicted cell abundances per spot for P1-P6. Black line represents median value, bar indicates interquartile range. **b**, Donut representation of predicted average abundance for the 10 most abundant cell types in My1-3 spots. Total (inside each donut) depicts average number of cells predicted across My1-3 spots. **c-l**, Predicted abundance of cell types in lesion core and T-rich regions (shown for patient P3).

**Extended Data Fig. 3:**
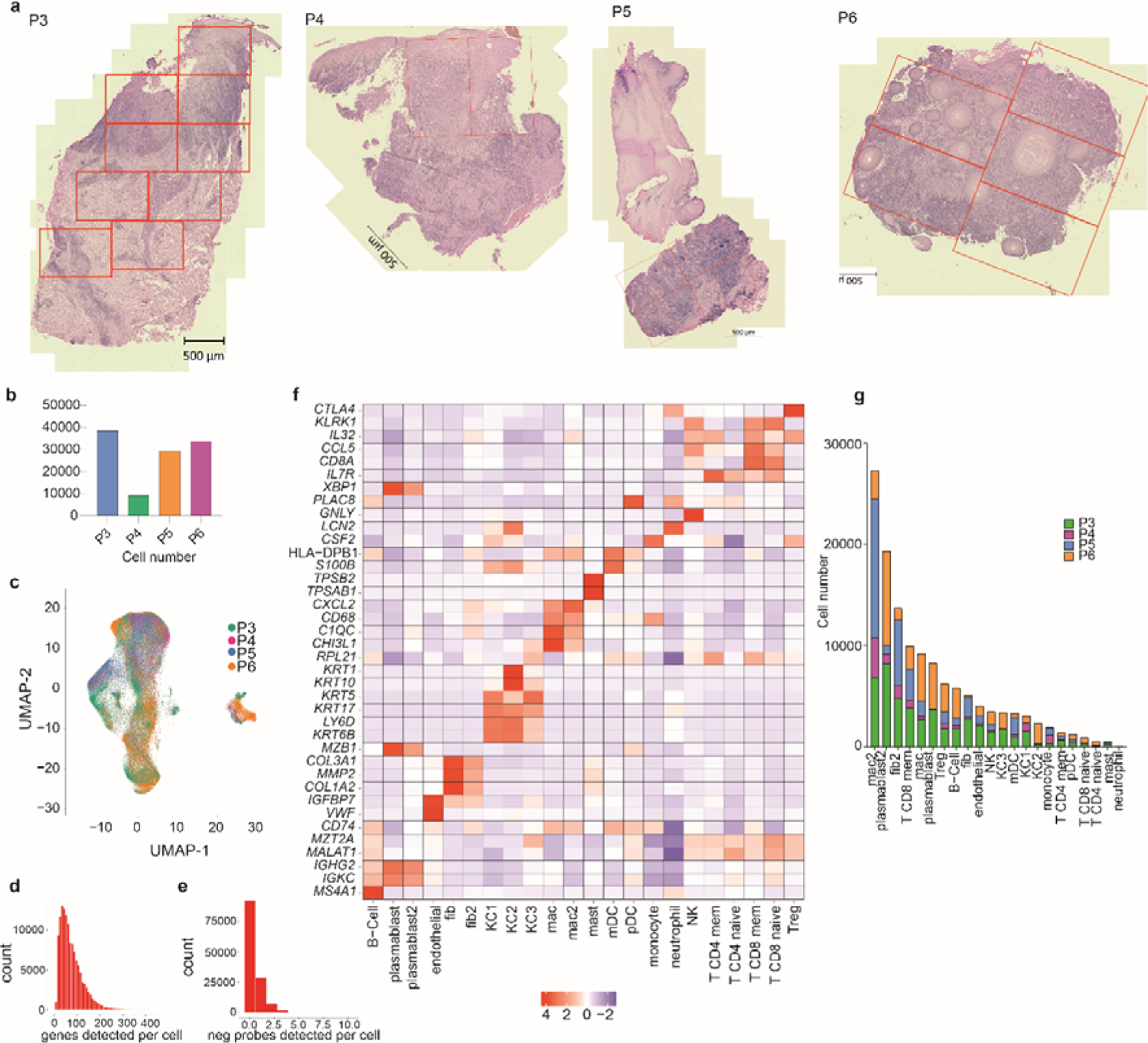
FOV positions, patient annotations, gene expression and cell number per cluster. **a,** H&E images for patients P3-P6 showing FOV analysed using Nanostring CosMx. Bar 500 µm **b,** Bar plots showing total cell numbers probed per patient. **c**, Cells were visualised in UMAP space and coloured by patient identity. **d-e**, Bar plots showing number of genes (**d)** and negative probes (**e**) detected per cell in the same dataset. **f**, Heat map showing top genes expressed by the identified cell types in all 4 patients. **g**, Stacked bar plot showing total cell counts across all patients for each population.

**Extended Data Fig. 4:**
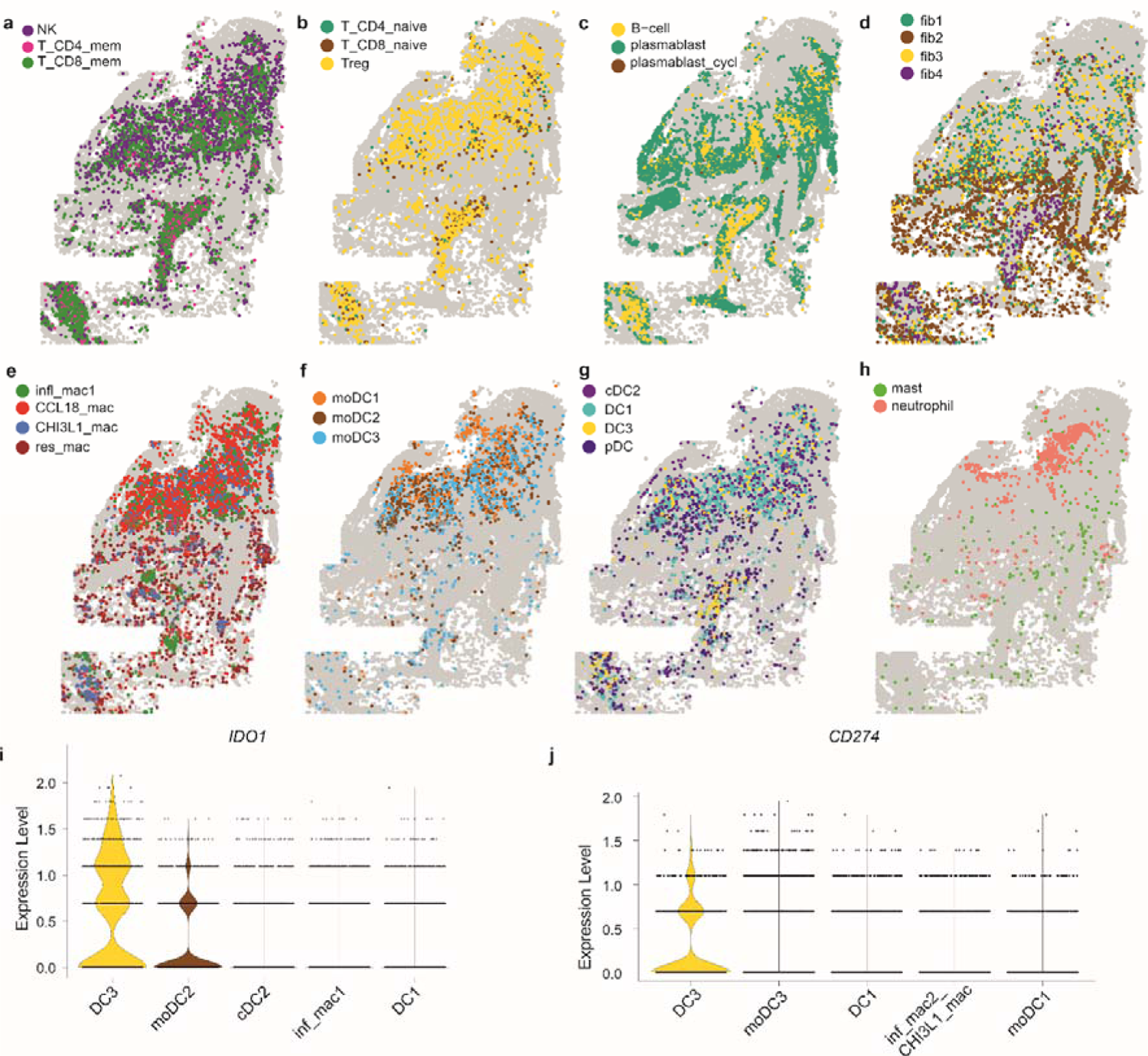
Spatial maps of cell types onto P3 section. **a-h,** Single-cell spatial mapping of indicated cell types for patient P3. **i-j,** Violin plots showing *IDO1* (**a**) and *CD274* (**b**) expression across DC3, moDC2, cDC2, infl_mac1 and DC1 cell types (n=4 patients). Average expression of *IDO1* and *CD274* was computed across all clusters and cell types that showed upper quartile expression were plotted.

**Extended Data Fig. 5:**
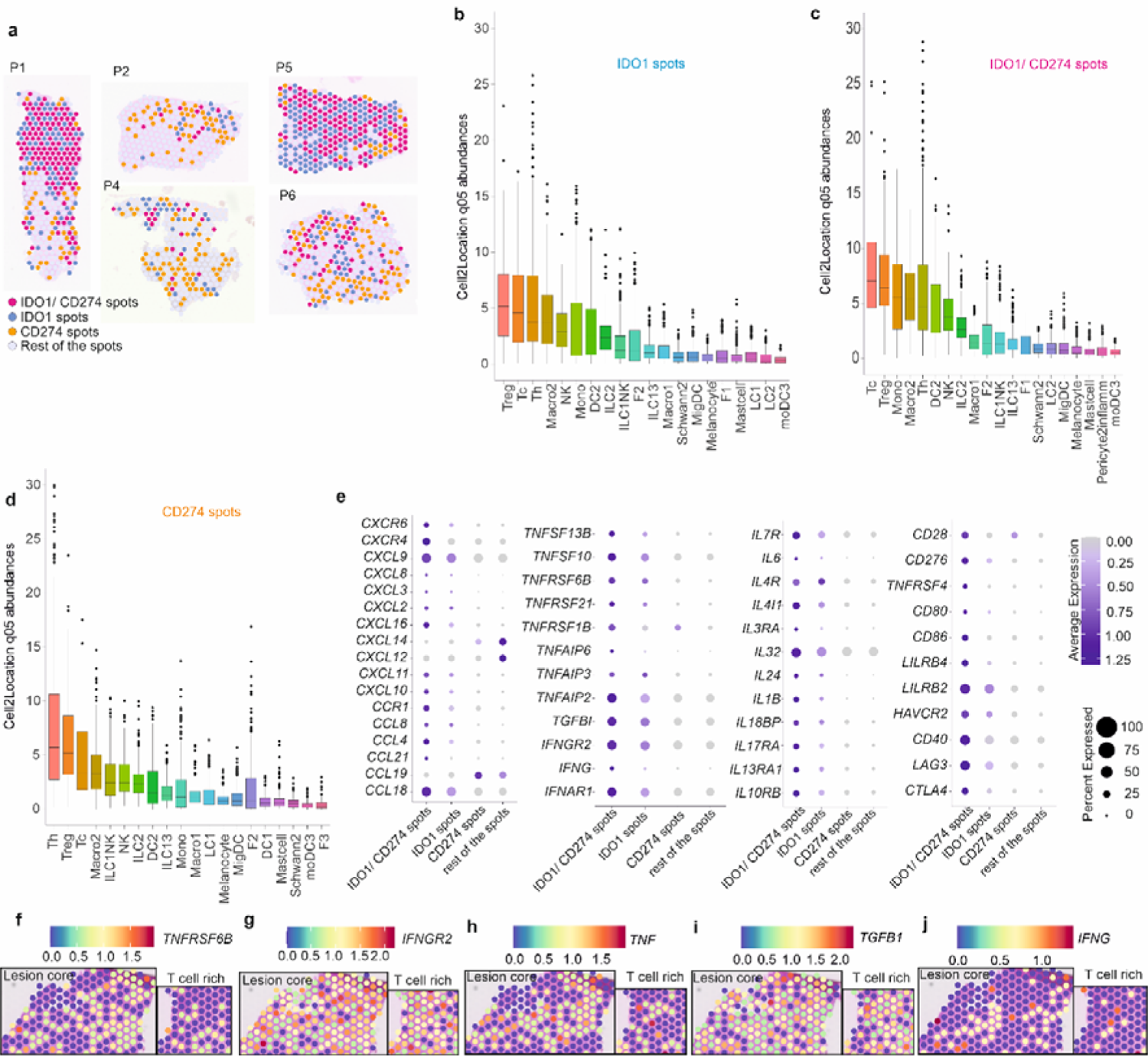
Characterisation of *IDO1*, *CD274* and *IDO1/ CD274* rich spots. **a,** Spatial feature plots for cytokines, chemokines and interleukins for patients P1, P2, P4-P6. **b-d,** Bar plot showing 5^th^ quantile top 20 predicted cell abundances for IDO and PD-L1 classes defined in (a) across all patient lesions (P1-P6). Black line represents median value, bar indicates interquartile range. **e**, Differential gene expression for selected genes across the grouping from (a). Log2Fold change cut off was set at 0.3 and p value >0.01. Data shown is from all 6 patients. **f-g,** Spatial feature plots for additional selected genes in lesion core and T cell rich area for patient P3.

**Extended Data Figure 6:**
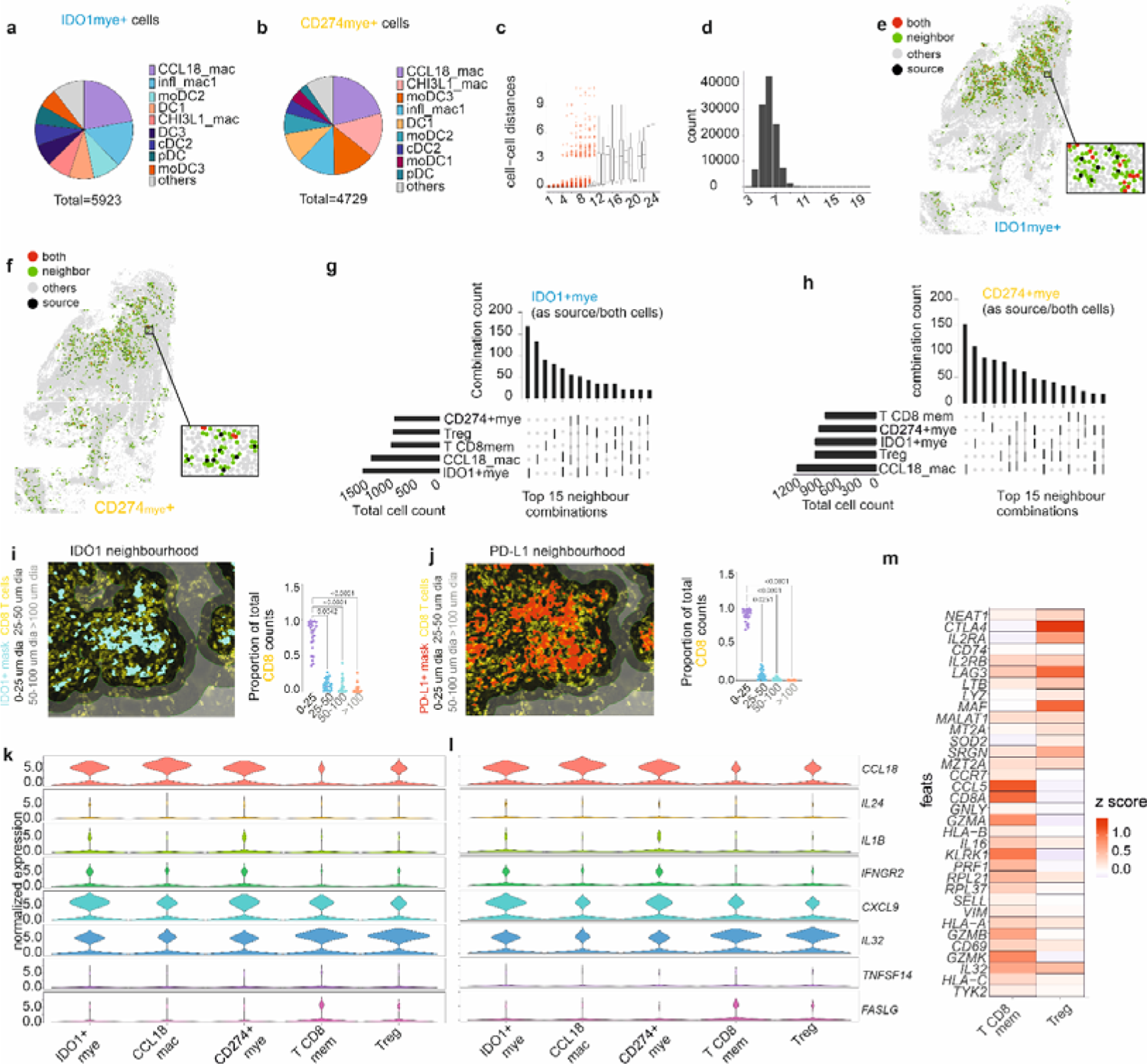
Phenotype and neighbours of IDO1^+^ and CD274^+^ myeloid cells. **a-b,** Pie chart for IDO1mye^+^ (**a**) and CD274mye^+^ (**b**) cells by myeloid subset (n=4 patients). **c**, Box and whiskers plots showing the cellular distances when a Delauney network is created for up to 22 immediate neighbours. Data for n=4 patients as in (a,b). **d,** Bar plot to show number of cells with between 3-22 immediate neighbours. **e**, **f.** Spatial plot of single cells for P3 coloured with “source” or “both” being either IDO1mye^+^ (e) or CD274mye^+^ (f). **g-h,** UpsetR plots for IDO1mye^+^ (g) or CD274mye^+^ (h) cells. 15 most frequent heterotypic interactions are shown from n= 5370 IDO1mye+ and n=4,308 CD274mye+neighbour source pairs, respectively. Connecting lines under the bar plot show neighbour combinations. We have also provided all the possible neighbour combinations in the source data file (**Supplementary Table 4**). Vertical bars represent total numbers of each combination while the horizontal plots indicate the total count of cell types as neighbours irrespective of the combination. **i-j,** Quantitative image analysis (n=23 patients) depicting proportions of CD8^+^ cells (yellow) present within 25,50, 100 µm and >100 µm distance (left panels) from IDO1^+^ (**i**) and PD-L1^+^ (**j**) cells. Cyan and red cell masks are drawn over IDO1 and PD-L1 positive cells respectively. Friedman’s one-way test with Dunn’s adjustment was applied for multiple comparisons. Adjusted p-value are given. **k-l,** Violin plots showing expression of *CCL18*, *IL24*, *IL1B*, *IFNGR2*, *CXCL9*, *IL32*, *TNFSF14* and *FASLG* in the top 5 neighbours of IDO1mye+ (**i)**, and CD274mye+(**j)** cells (n=4 patients). **m,** Heat map showing phenotype of T CD8 memory and Treg cells. Scale shows gini coefficient z scores (n=4 patients).

**Extended Data Fig. 7:**
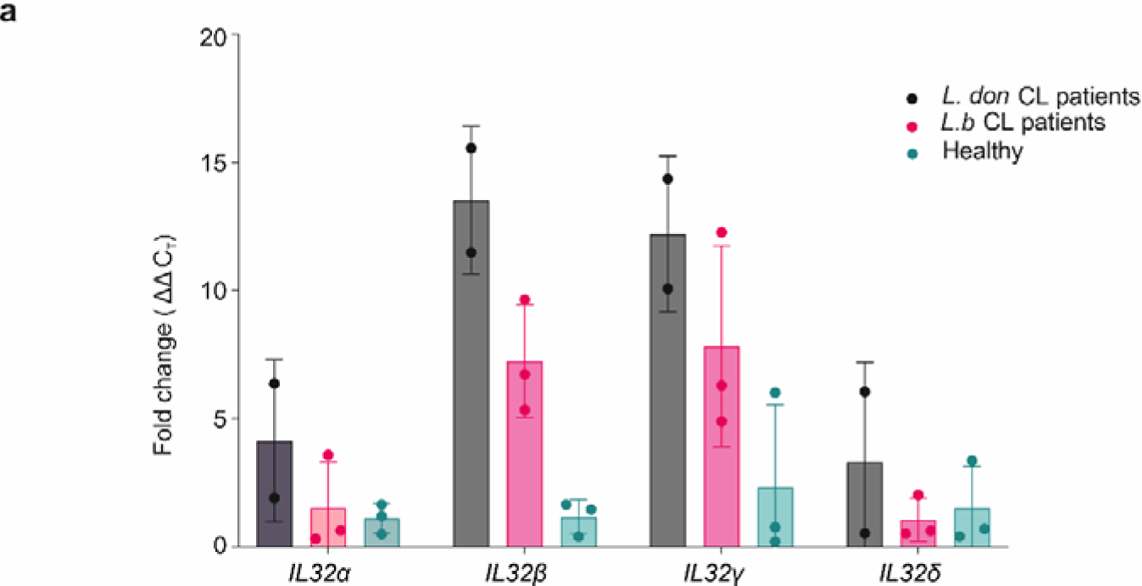
Isoforms of *IL32* in CL patients. **a,** Fold change of *IL32* α, *ß*, γ and δ isoforms with respect to GAPDH and healthy skin in Sri Lankan CL patients (n=2) and *L*.(*V.*) braziliensis CL patients. Bars show mean +/- standard deviation.

**Extended Data Fig. 8:**
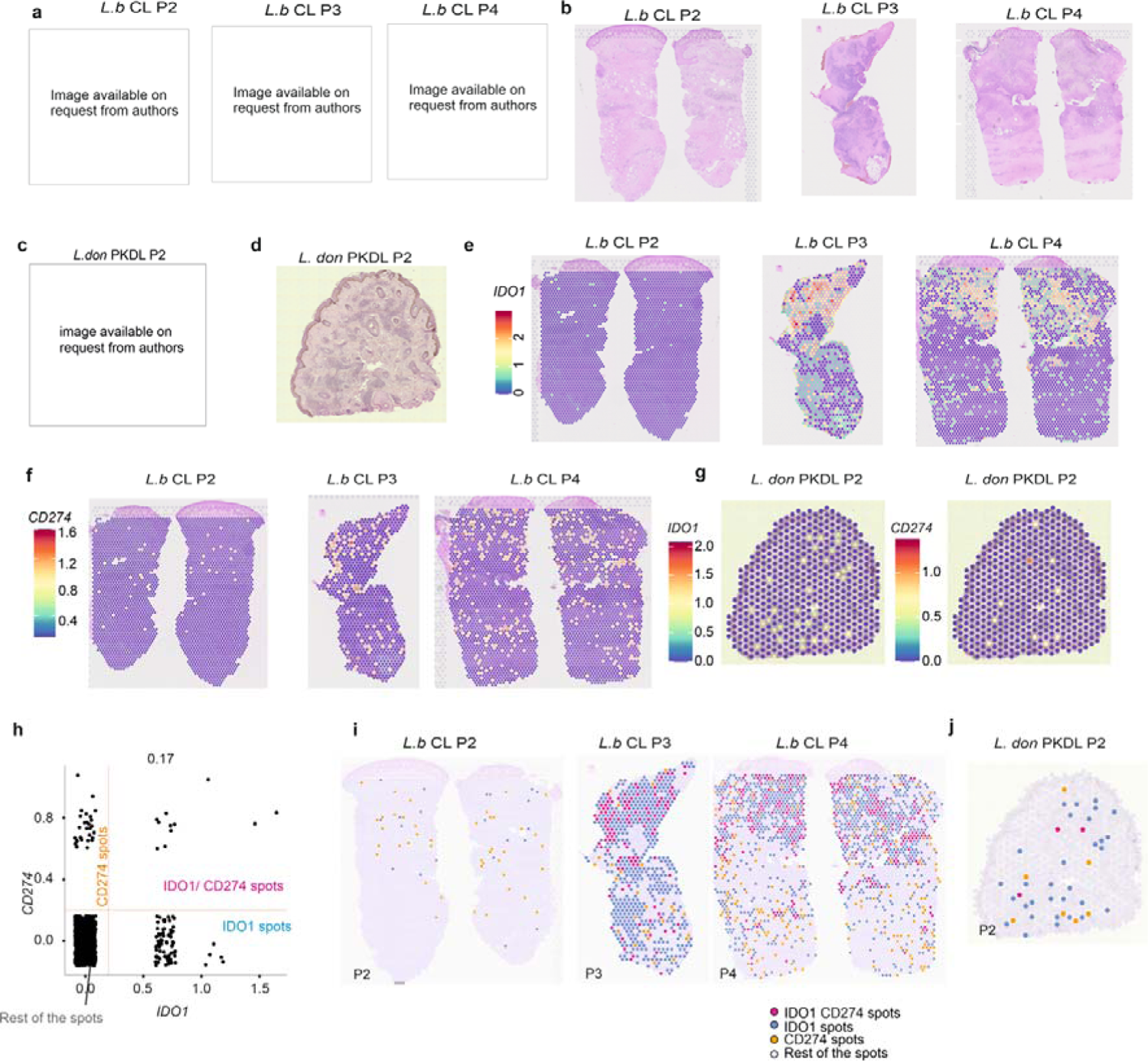
*IDO1*+ and *CD274*+ microenvironments in *L. braziliensis* and *L. donovani* infected CL and PKDL lesions respectively. **a,** CL lesions caused by *L. braziliensis.* Images for patients *L.b*_P2-P4. **b,** Matched H&E images of 5μm sections of lesions in (**a**). Mild inflammatory infiltrate and extensive fibrosis was seen in *L.b*._P2. **c-d,** Macular PKDL lesion caused by *L. donovani.* Image for patient PKDL_P2 (**c**) and corresponding histology shown in an H&E image **(d)**. **e-f,** Spatial plots showing gene expression for *IDO1* (e) and *CD274* (f) for patients *L. b.*_P2-P4. P2 showed very limited expression of *IDO1* and *CD274* with no overlap between the two expression patterns. **g,** Spatial plots showing the gene expression of *IDO1* (**c)** and *CD274* **(d)** for macular *L. don* PKDL_P2. **h,** Scatter plot showing *CD274* and *IDO1* expression across all spots from *L. don* PKDL_P1 and P2, with thresholds at y=0.2 and x=0.2. **i-j,** Spatial plots coloured by class for *L.b*_P2-4 **(i)** and PKDL_P2 (**j**).

**Extended Data Fig. 9:**
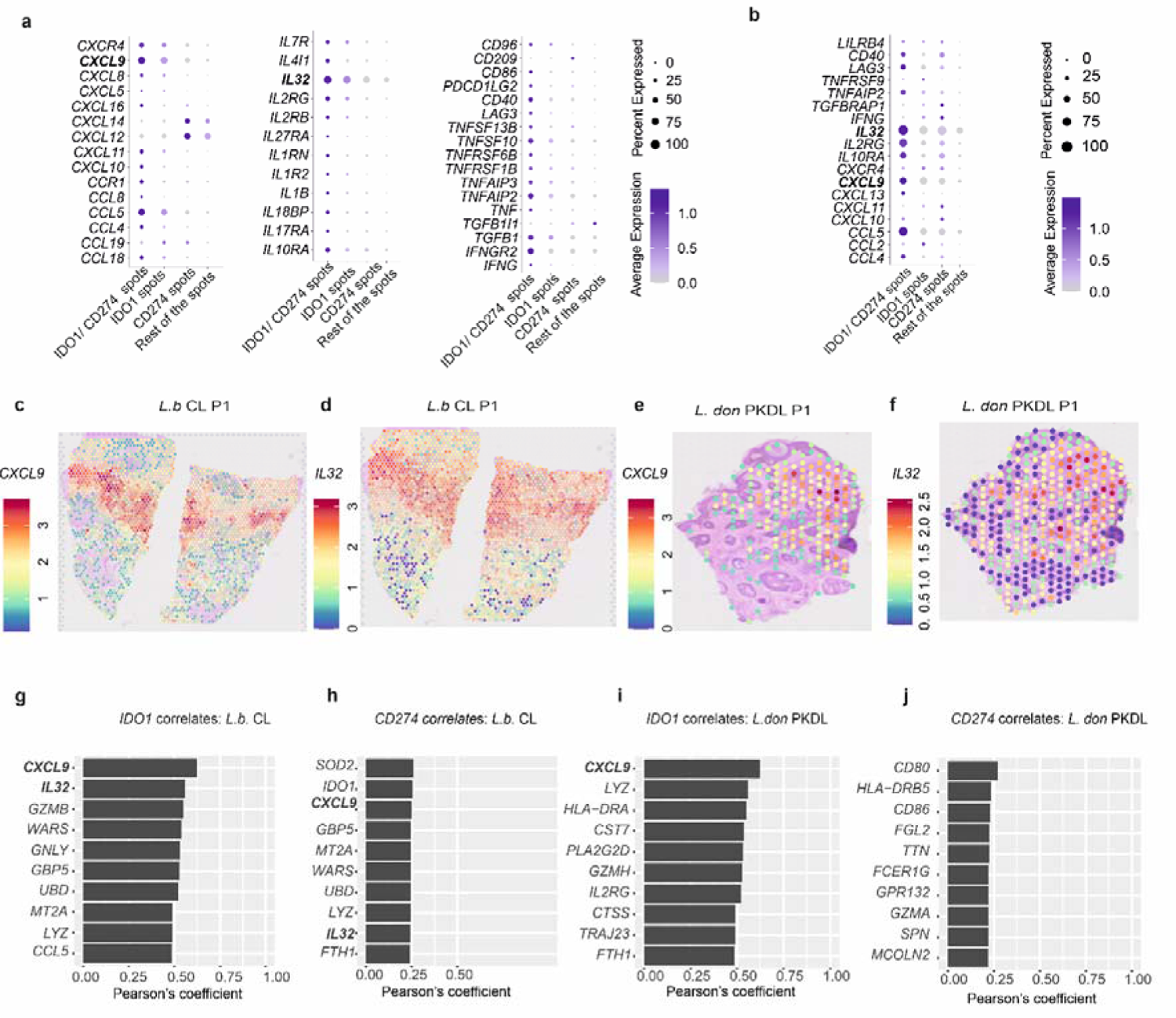
Phenotyping *IDO1*^+^ and *CD274*^+^ microenvironments in *L. braziliensis* CL and *L. donovani* PKDL and lesions. **a,** Dot plot showing differentially expressed cytokines, chemokines, interleukins, TNF and interferon related and immune checkpoint markers between IDO1 and CD274 classes in *L.b.* CL patients (n=4; a) and PKDL patients (n=2, b). Cut off was p<0.05, Log2Fold = 0.25. **c-f,** Spatial plots showing gene expression for *CXCL9* and *IL32* in *L.b*_P1 and PKDL_ P1. **g-j,** Genes correlating with *IDO1* (**g,i),** and *CD274* (**h, j)** from all *L. b*. CL patients (g,h) and PKDL patients (i-j). Pearson’s correlation coefficients are plotted in decreasing order (p<0.05).

