## Supplementary Tables 1-3 for "IL-32 producing CD8^+^ memory T cells and Tregs define the IDO1 / PD-L1 niche in human cutaneous leishmaniasis skin lesions"

**Table 1 : Demographics of Sri Lankan cutaneous leishmaniasis (CL; *L. donovani* ) patients**

| PID | Age range | Gender | Duration of lesion in months | Comments | Amastigote grade | SSS | PCR | Amastin (by RNA-FISH) | Treatment response at the end of treatment | total SSG doses for cure |
|---|---|---|---|---|---|---|---|---|---|---|
| P1 | >60 | M | 2 | Primary lesion on right arm. Three more on right arm, chest, and back | 3+ | Positive | Positive | Positive | Complete cure | 10 |
| P2 | 46-60 | F | 8 | Primary lesion on left scapular area | 1+ | Positive | Positive | Positive | Complete cure | 17 |
| P3 | 46-60 | M | 5 | Primary lesion on right shoulder, another lesion on right arm | 2+ | Positive | Positive | Positive | Complete cure | 7 |
| P4 | 31-45 | M | 9 | Primary lesion on left arm | 2+ | Positive | Positive | Positive | Complete cure | 21 |
| P5 | 0-30 | M | 25 | Primary lesion on left forearm | 0 | Negative | Positive | Positive | Complete cure | 7 |
| P6 | 46-60 | F | 3 | Primary lesion on right upper neck, another on right shoulder | 5+ | Positive | Positive | Positive | Not completely healed. Still on treatment at 6 mo FU | 29 |
| P7 | 0-30 | M | 3 | Primary lesion on jaw . Three more on chin and forearm. | 5+ | Positive | Positive | Negative | Complete cure | 5 |
| P8 | 46-60 | M | 37 | Primary lesion on right forearm | 0 | Negative | Positive (Faintly) | Negative | Complete cure | 12 |
| P9 | 46-60 | M | 4 | Primary lesion on left arm | 0 | Negative | Positive (Faintly) | Negative | Complete cure | 10 |
| P10 | 31-45 | M | 13 | Primary lesion on left elbow | 1+ | Positive | Positive | Negative | Complete cure | 6 |
| P11 | 31-45 | F | 7 | Primary lesion on right shoulder | 1+ | Positive | Positive (Faintly) | Negative | Complete cure | 6 |
| P12 | 31-45 | M | 7 | Primary lesion on left hand | 4+ | Positive | Positive | Positive | Complete cure | 8 |
| P13 | 31-45 | M | 3 | Primary lesion on right upper back, another on right hand. | 2+ | Positive | Positive | Negative | Complete cure | 9 |
| P14 | 46-60 | M | 13 | Primary lesion on left forearm | 2+ | Positive | Positive | Negative | Complete cure | 16 |
| P15 | 31-45 | F | 2 | Primary lesion on left leg, another on right ankle | 2+ | Positive | Positive | Negative | Complete cure | 12 |
| P16 | 46-60 | M | 6 | Primary lesion on right forearm | 5+ | Positive | Positive | Negative | Complete cure | 13 |
| P17 | 31-45 | M | 8 | Primary lesion on right forearm | 0 | Negative | Positive | Negative | Complete cure | 12 |
| P18 | 31-45 | M | 6 | Primary lesion on right forearm | 0 | Negative | Positive | Negative | Complete cure | 9 |
| P19 | 46-60 | M | 4 | Primary lesion on back of the neck | 1+ | Positive | Positive | Negative | Complete cure | 6 |
| P20 | 46-60 | M | 4 | Primary lesion on right arm, another on right back abdomen | 0 | Negative | Positive (Faintly) | Negative | Complete cure | 15 |
| P21 | >60 | M | 3 | Primary lesion on left leg, another on left leg lower anterior | 0 | Negative | Positive | Negative | Complete cure | 10 |
| P22 | 31-45 | F | 3 | Primary lesion on left arm | 2+ | Positive | Positive | Negative | Complete cure | 9 |
| P23 | >60 | M | 2 | Primary lesion on left leg and another on left cheek | 2+ | Positive | Positive | Negative | Complete cure | 4 |
| P24 | 46-60 | M | 5 | Primary lesion on right arm and two more on right shoulder and left forearm | 2+ | Positive | Positive | Negative | Complete cure | 26 |
| P25 | 0-30 | M | 4 | Primary lesion on left forearm | 2+ | Positive | Positive | Negative | Complete cure | 12 |

**Table 2 : Demographics of Brazillian cutaneous leishmaniasis (CL; *L. braziliensis* ) patients**

| PID | Age range | Gender | Lesion size mm2 | Lesion duration (in months) | DX PCR | DX IIF | DX ELISA | Treatment; route | Duration of therapy | t/t visual score BT | t/t visual score 1 week | t/t visual score 2 week | t/t visual score 3 week | t/t visual score 4 week | t/t visual score 3 months | t/t visual score 6 months |
|---|---|---|---|---|---|---|---|---|---|---|---|---|---|---|---|---|
| P1 | 46-60 | F | 2400 | 3 months | PCR + | - | + | liposomal amphotericin B; systemic | 10 days | 10 | 10 | 10 | 10 | 9 | 5 | 2 |
| P2 | 31-45 | M | 1225 | 2 months | PCR + | + | + | Glucantime (meglumine antimoniate); systemic | 21 days | 10 | 10 | 10 | 5 | 3 | 0 | 0 |
| P3 | 31-45 | M | 5525 | 6 months | PCR + | + | + | Glucantime (meglumine antimoniate); systemic | 20 days | 10 | 10 | 8 | 8 | 8 | 0 | 0 |
| P4 | >60 | M | 750 | 6 months | PCR+ | - | + | Glucantime (meglumine antimoniate); intralesional | 4 weeks | 10 | 10 | 8 | 8 | 8 | 5 | 0 |

**Table 3 : Demographics of Indian  post kala-azar leishmniasis (PKDL; *L. donovani* )  patients**

| PID | Age range | Gender | lesion duration | History of VL | Year of VL | Treatment during VL | Year of PKDL | Lag period (in years) | Lesion type | DX ITS-1 PCR |
|---|---|---|---|---|---|---|---|---|---|---|
| P1 | 0-30 | M | 3 years | Yes | 2010 | SAG | 2016 | 6 | Polymorphic | Positive |
| P2 | 31-45 | M | 8 years | Yes | 1989 | SAG | 2013 | 24 | Macular | Positive |
