## Supplementary Figures 1-3 for "IL-32 producing CD8^+^ memory T cells and Tregs define the IDO1 / PD-L1 niche in human cutaneous leishmaniasis skin lesions"

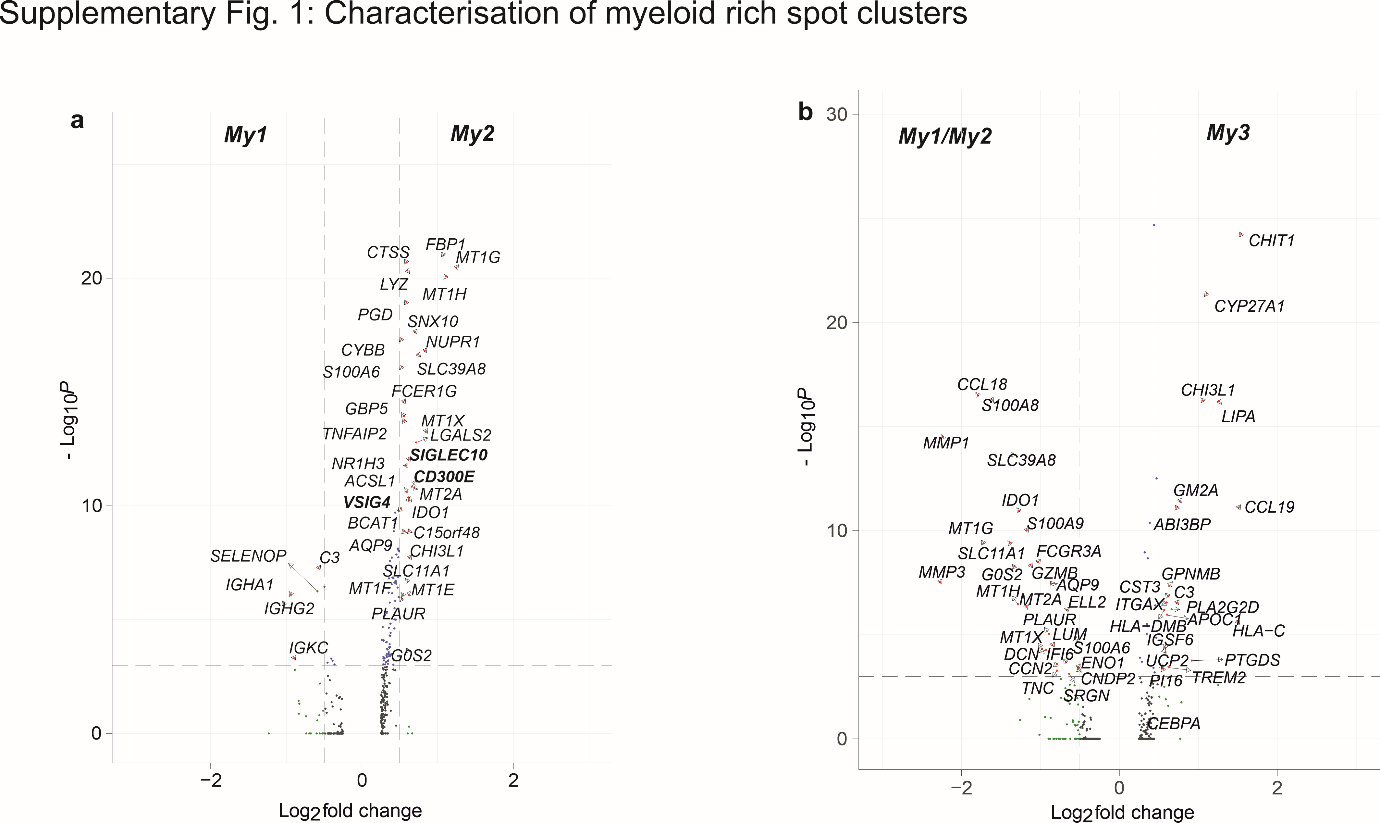

**Supplementary Fig. 1:** **Characterisation of Myeloid rich spot clusters**

**a-b,** Volcano plot showing differentially regulated genes in My1 vs. My2 spots (a) and inn My1/My2 vs. My3 spots (b). p-values are thresholded at -Log_10_P value of 3 and Log2Fold change of -0.5 and 0.5.

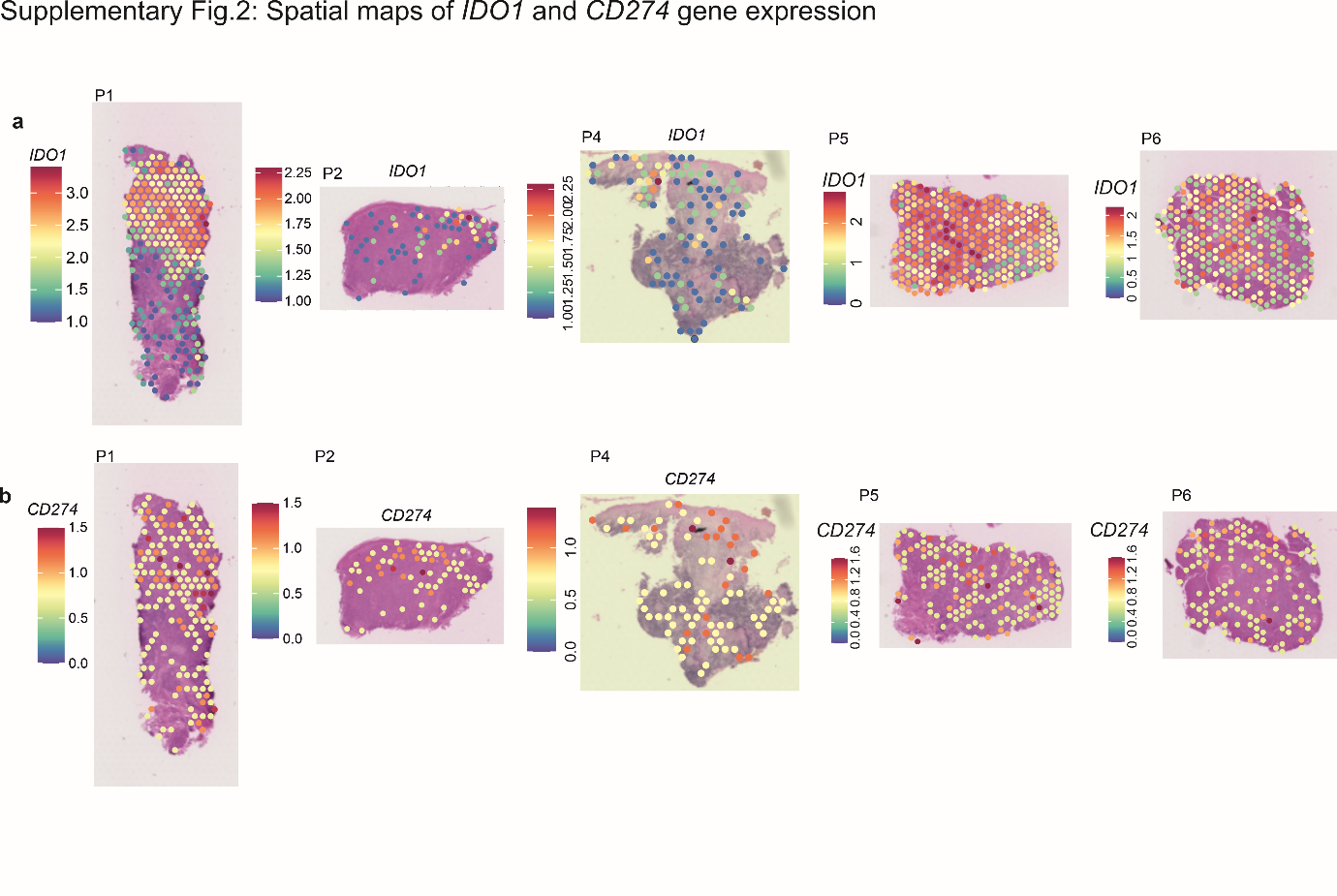

**Supplementary Fig. 2: Spatial maps of *IDO1* and *CD274* gene expression**

**a-b,** Spatial plot showing expression of *IDO1* (**a)** and *CD274* (**b**) for patients P1, P2, P4, P5 and P6.

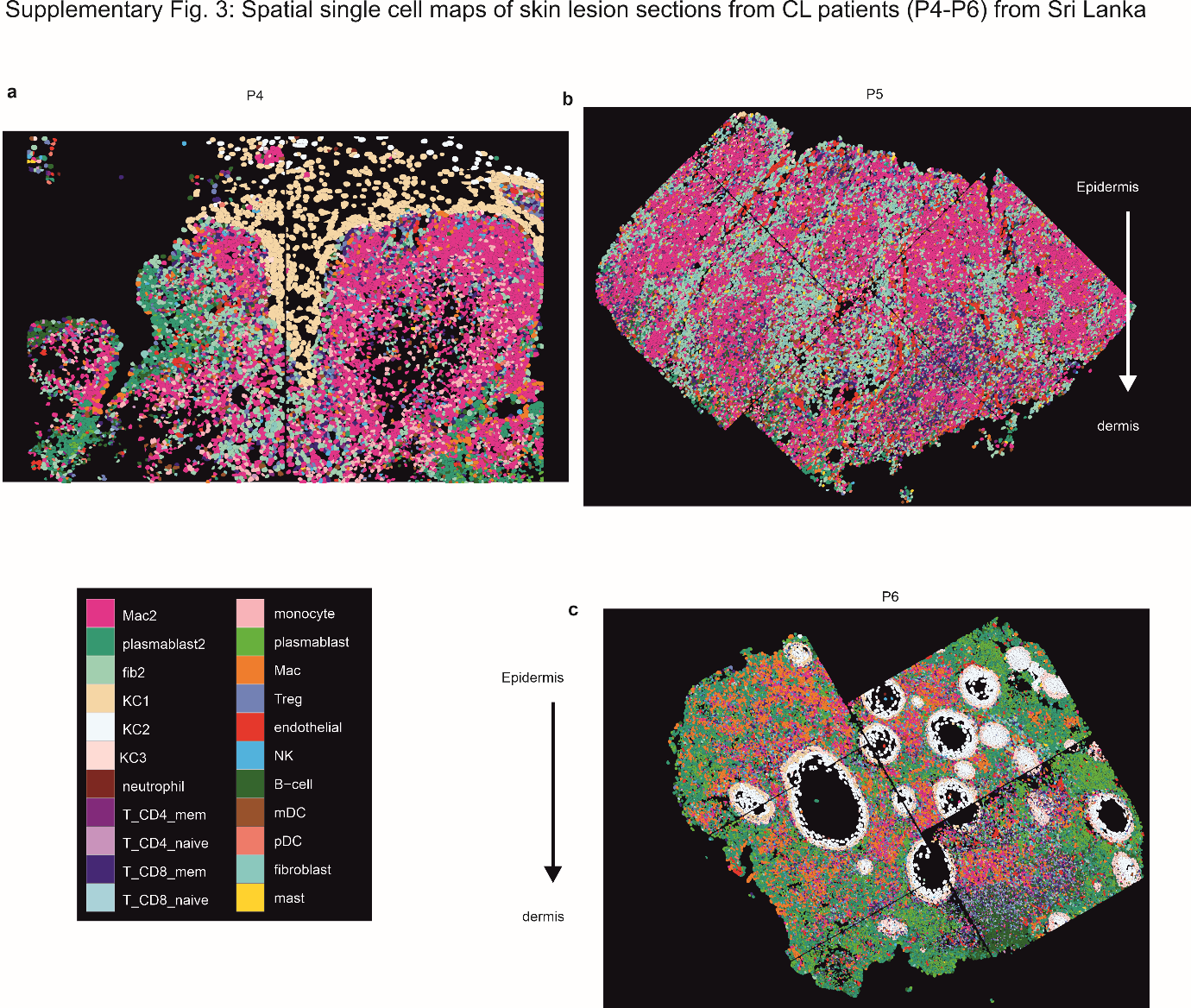

**Supplementary Fig. 3: Single cell spatial maps of skin lesions from CL patients in Sri Lanka**

**a-c,** Single-cell spatial maps of skin lesions for patients P4 (**a**), P5 (**b**) and P6 (**c**) labelled by their cell types as identified by transcriptomic profile.
